# Common anti-platelet therapies modulate procoagulant phospholipids in arterial disease

**DOI:** 10.1101/2022.11.03.22280948

**Authors:** Majd B Protty, Victoria J Tyrrell, Ali A Hajeyah, Bethan Morgan, Yong Li, Anirban Choudhury, Rito Mitra, David Bosanquet, Alastair W Poole, Zaheer Yousef, Peter W Collins, Valerie B O’Donnell

## Abstract

Enzymatically oxygenated phospholipids (eoxPL) formed by lipoxygenases (LOX) and cyclooxygenase (COX) in platelets and leukocytes are pro-coagulant in multiple model systems. However, their generation in arterial thrombotic disease, and how their levels are modulated by common therapies is unknown. Here, eoxPL were first characterized in isolated platelets and leukocytes from an arterial vascular disease cohort, a healthy cohort administered low dose aspirin, and from retrieved human arterial thrombi. In both cohorts, aspirin reduced platelet COX-1-derived eoxPL, while elevating diacyl 12-LOX-derived eoxPL in males, through enhanced Lands’ cycle esterification. Conversely, P2Y12 inhibition reduced 12-LOX-derived eoxPL in leukocytes. Complex aspirin-dependent gender and seasonal effects on platelet eoxPL were seen in healthy subjects. Limb or coronary (STEMI) thrombi showed a platelet eoxPL signature while carotid thrombi had a white cell profile. Mice genetically lacking leukocyte 12/15-LOX, which are deficient in eoxPL, generated smaller carotid thrombi in vivo. In summary, pro-coagulant eoxPL generation is altered in human arterial vascular disease by commonly used cardiovascular therapies. These changes to the phospholipid composition of blood cells in humans at risk of thrombotic events may be clinically significant where the pro-coagulant membrane plays a central but poorly understood role in driving elevated thrombotic risk.

**Key Points:**

- eoxPL generation is altered in health and arterial vascular disease by aspirin or P2Y12 inhibitors, and shows gender and seasonal variation.
- Aspirin regulates eoxPL by inhibiting cyclooxygenase and modulating Lands’ cycle.
- The eoxPL profile of human arterial thrombi identifies platelet and leukocyte involvement.
- Mice deficient in LOX-derived eoxPL form smaller arterial thrombi *in vivo*.

## Introduction

Coagulation factor activity, to form an occlusive thrombus, requires the presence of an electronegative pro-coagulant membrane, traditionally considered to comprise phosphatidylserine (PS) and phosphatidylethanolamine (PE). In this model, resting cells maintain most PS and PE on the inner leaflet of their plasma membrane, but on activation (e.g. thrombin activation of platelets) externalize up to 7 - 10 % via scramblase activity^1, 2^. Through associating with calcium and supported by PE, the PS headgroup facilitates the binding and activity of coagulation factors, ultimately allowing formation of thrombin from prothrombin^3–6^. In recent years, a central role for “enzymatically-oxidized phospholipids” (eoxPL), in particular hydroxyeicosatetraenoic acids (HETEs) attached to PE or phosphatidylcholine (PE) in coagulation has been proposed^7, 8^. HETE-PL are acutely generated on activation of platelets or leukocytes and a proportion become externalized on the outer membrane^5, 9–12^. Extensive mechanistic studies demonstrated that eoxPL act in concert to support the action of PS on the membrane surface through mediating biophysical changes^13^. Calcium binding is increased and the accessibility of the PS headgroup to interact with factors appears to be enhanced, supporting increased thrombin generation^8, 13^. The role of eoxPL in driving coagulation *in vivo* has only been examined in a small number of studies so far. These include venous thrombosis in mouse models and anti-phospholipid syndrome in humans^8, 14^. Mice lacking the lipoxygenase (LOX) enzymes that generate eoxPL (either *Alox15* or *Alox12*) generate smaller venous clots and bleed longer in challenge models^8, 14^. Recently, a role for the LOX enzymes that generate eoxPL in driving abdominal aortic aneurysm (AAA) in mice was demonstrated, and eoxPL were detected in human and murine AAA lesions^15^.

Up to now, the generation and action of eoxPL in arterial thrombosis, particularly in human coronary disease, has not been studied, and their role in regulating coagulation in this disease is unknown. Acute coronary syndrome (ACS) is a common manifestation of arterial thrombosis. It leads to ischemic heart disease and is associated with high rates of mortality, recurrent infarction and other complications^16–20^. Along with this, inflammation is a central feature of ACS, where plaque rupture occurs along with recruitment of platelets^21, 22^, leukocytes^23^ and tissue factor upregulation^24–28^. During thrombo-inflammation, coagulation leads to an occlusive arterial thrombus^29^. Despite the use of anti-platelet therapy, rates of stroke, myocardial infarction and cardiovascular death remain relatively high during the first year following diagnosis^30, 31^. This suggests that there are other factors at play beyond platelets^30–32^. One potential player is the coagulation cascade resulting in formation of thrombin (Factor IIa)^33^, driven by pro-coagulant membranes, including those containing eoxPL.

Here, to test their potential involvement in arterial disease, we determine the eoxPL profile on the membranes of platelets, leukocytes and extracellular vesicles (EVs) in ACS, and following aspirin supplementation. In a healthy cohort, we examine the impact of aspirin, gender, seasonality and cell activation on eoxPL generation. The molecular species of eoxPL in human arterial thrombi are characterized, and the role of *Alox15* in arterial thrombosis determined in a mouse model.

## Methods

### Study Participants

*Healthy cohort:* Healthy volunteers (n = 28) were recruited from the workplace (14 male, 14 female; age range 20-50 years). Ethical approval, which included informed consent, was from Cardiff University, Schools of Medicine and Dentistry Research Ethics Committee (SMREC16/02), and the study is registered as NCT05604118 (ClinicalTrials.org). Following a 2-week period free from non-steroidal anti-inflammatory drugs (NSAIDs), peripheral blood was obtained and platelets isolated as outlined below. Participants were commenced on aspirin 75 mg once daily for seven days and then provided a repeat blood sample. Two (one male, one female) were unable to provide a sample post aspirin treatment. Following a 2-month period, a subset of subjects (7 male, 7 female) returned and provided repeat samples pre- and post-aspirin, and again a third time 2 months later.

*Clinical cohort:* Participants were recruited from Cardiff University and Cardiff and Vale University Health Boards. Ethical approval was from Health and Care Research Wales (HCRW, IRAS 243701; REC reference 18/YH/0502). Study groups of at least 20 were aimed for based on a previous study in venous thrombosis^8^. Age and gender-matched individuals were recruited into one of the following four groups: *(i) Acute Coronary Syndrome (ACS)*: Participants were identified on in-patient cardiology wards using diagnostic tests (ischemic ECG changes, raised troponin level above normal laboratory defined range) and clinical assessment by the cardiology team. All were recruited within 48 hours of the index event prior to any revascularization/angioplasty. *(ii) Significant coronary artery disease (CAD):* Patients attending for an elective coronary angiogram to assess for symptoms of stable angina in the absence of a history of acute coronary syndrome were recruited. Coronary angiography demonstrated lesions requiring revascularization on anatomical/physiological criteria as defined by guidelines from the European society for cardiology (ESC, 2018)^34^. *(iii) Risk-factor controls with no significant CAD (RF):* This group includes patients attending for a diagnostic coronary angiogram with any risk factors for ischemic heart disease (a clinical diagnosis of hypertension requiring therapy, diabetes types 1 or 2, hypercholesterolemia [total cholesterol > 6 mmol/L], smoking, chronic kidney disease stage 3 or more, or combination thereof) but whose coronary angiogram demonstrates no significant coronary artery disease, defined as not requiring revascularization on anatomical/physiological criteria as per the ESC 2018 guidelines^34^. *(iv) Healthy controls (HC):* Participants had no history of ischemic heart disease or its risk factors, were never-smokers, and were not on anti-platelet agents, anti-coagulants, anti-hypertensives or statins. They were identified from the workplace or were volunteers from partner studies such as ‘HealthWise Wales’^35^. Clinical characteristics are in Supplementary Table 1. Inclusion criteria were aged 18 yrs or older, acute coronary syndrome in ACS group, and no history of ACS in the others. Exclusion criteria were: diagnosis of infective endocarditis or atrial fibrillation, or inability to consent to study. Here, 90 patients were recruited: HC, n = 24, RF, n = 23, CAD, n = 19 and ACS, n = 24. Blood samples were collected by peripheral venepuncture as outlined below, by one individual and all samples were transferred to the laboratory within 10 min.

### Platelet isolation

Whole blood was taken from the antecubital vein using a 21G butterfly into a 50 ml syringe containing acidified citrate dextrose (ACD; 85 mM trisodium citrate, 65 mM citric acid, 100 mM glucose, pH 5.0) at a ratio of 8.1 parts whole blood to 1.9 parts ACD, as described previously^8^, and centrifuged at 250 × *g* for 10 min at 20 °C. The platelet-rich plasma was collected and centrifuged at 1000 × *g* for 8 min at 20 °C. Platelet poor plasma was removed and retained for extracellular vesicle isolation. The platelet pellet was resuspended in Tyrode’s buffer (134 mM NaCl, 12 mM NaHCO_3_, 2.9 mM KCl, 0.34 mM Na_2_HPO_4_, 1.0 mM MgCl_2_, 10 mM HEPES, 5 mM glucose, pH 7.4) containing ACD (9:1, v/v). The platelets were washed by centrifuging at 1000 × *g* for 8 min at 20 °C then resuspended in Tyrode’s buffer at a concentration of 2 × 10^8^·ml^−1^. Platelets were activated at 37 °C in the presence of 1 mM CaCl_2_, 0.2 unit·ml^−1^ thrombin (Sigma Aldrich) for 30 min with occasional inversion.

### Leukocyte isolation

Leukocytes were isolated from 20 ml citrate-anticoagulated whole blood as described previously^8^. Briefly, 20 ml of blood was mixed with 4 ml of 2 % citrate and 4 ml of Hetasep (Stem Cell Technologies) and allowed to sediment for 45 minutes. The upper plasma layer was recovered and centrifuged at 250 g for 10 min at 4 °C. The pellet was resuspended in ice-cold 0.4 % trisodium citrate/PBS and centrifuged at 250 g for 5 min at 4 °C. Erythrocytes were removed by hypotonic lysis (0.2 % hypotonic saline). Leukocytes were resuspended in Krebs buffer (100 mM NaCl, 48 mM HEPES, 5 mM KCl, 1 mM sodium dihydrogen orthophosphate dihydrate and 2 mM glucose) at 4 x 10^6^/ml. For activation, 4 x 10^6^ leukocytes were incubated at 37 °C with 10 μM A23187 and 1 mM CaCl_2_, for 30 min with occasional inversion, prior to lipid extraction.

### Extracellular vesicle (EV) isolation

Methods were adapted from recent literature and guidelines^36, 37^. Platelet poor plasma was generated above was centrifuged at 1000 x g for 10 min at 20 °C to generate platelet-free plasma (PFP). 1 ml of this was snap frozen on dry ice and stored at -80 °C for EV quantification at a later date see below). For each donor plasma, 6 x 1 ml PFP aliquots were centrifuged at 16,000 g for 30 min at 20 °C. 750 μl was removed from each aliquot, and 750 μl of modified Tyrode’s buffer was added to the pellet, which was gently resuspended using a pipette. Following a second centrifugation at 16,000 g for 30 min at 20 °, 950 μl was removed. 50 μl modified Tyrode’s buffer was added to the pellet to gently resuspend and recover the EV-rich fraction. The EV fractions were pooled to generate one isolate per donor. Of this, 250 μl was used for lipidomics, and 3 x 20 μl for prothrombinase assays. Thus, the final samples were 10-fold concentrated from the original plasma EV concentration.

### Extracellular vesicle (EV) quantification

EV quantification was performed by thawing one aliquot of PFP per patient, of which 500 μL was passed through size-exclusion chromatography iZON qEV columns (Izon Science Ltd, UK) to recover particles and vesicles between 70 nm to 1000 nm in diameter. The eluting EV-rich fractions were collected and analysed using nanoparticle tracking on a NanoSight 300 (Malvern, UK) equipped with a sensitive sCMOS camera and a 488 nm blue laser, to generate an EV count and size distribution for all participants’ plasmas.

### Human Thrombi

The study took place between Cardiff University, Cardiff and Vale University Health Board (CVUHB) and Aneurin Bevan University Health Board (ABUHB). Ethical approval was from Health and Care Research Wales (HCRW, IRAS 243701; REC reference 18/YH/0502). Patients were identified by the collaborating vascular/cardiology teams from their operating patient lists. There were no changes to the standard operating procedure and no additional surgical/interventional steps were carried out. Some of these procedures were time-critical, and ethical approvals allowed the research team to seek retrospective consent in the following 48 hours after the emergency procedure. This was in the form of written informed consent which also included permission to record the participants medical history from hospital notes. If consent was declined, stored samples were discarded as per local procedures. Inclusion criteria were 18 yrs of age or older and undergoing vascular surgery or percutaneous transluminal intervention to treat arterial thrombotic disorders, where there was a clinical indication to remove and discard diseased tissue. Exclusion criteria was inability to consent within specified time frame. Thrombi were obtained from human patients as follows: *(i) Coronary thrombi* were collected from patients presenting with ST-elevation myocardial infarction (STEMI) to the Cardiology Department in Cardiff and Vale University Health Board and underwent emergent percutaneous coronary intervention (PCI). Patients were diagnosed on arrival based on electrocardiographic features of ST-segment elevation in more than 2 leads corresponding to a myocardial muscle territory. Following this, patients were immediately prepared for coronary angioplasty. A thrombus aspiration catheter (Export Advance^TM^, Medtronic, Ireland) was advanced through the peripheral vascular access (radial artery) to the coronary artery and the thrombus retrieved. This was washed with saline using a cell sieve, transferred to a sterile cryovial and immediately snap frozen on dry ice prior to transfer to -80°C. *(ii) Carotid thrombi* were collected from patients at the Vascular Department at ABUHB identified from routine operating lists, who were undergoing carotid endarterectomy, typically following an ischemic stroke/transient ischemic attack with evidence of >50% stenosis of the appropriate internal carotid artery on duplex ultrasonography. *(ii) Limb thrombi* were taken from patients with acute limb ischemia requiring either embolectomy, thrombectomy, arterial bypass or amputation. For carotid and limb clots, the clot was retrieved using standard open surgical techniques, placed into cryovials and immediately snap frozen on dry ice prior to transfer to -80 °C freezer for storage. Supplementary Tables 2 and 3 show clinical demographics of patients including biochemical and hematological characteristics.

### Murine model of carotid artery injury

Female mice (12 weeks old C57/B6/J) were purchased from Charles River UK (Margate, UK), while *Alox15^-/-^* mice were bred in house (F11, C57BL/6J) in isolators at Cardiff University. All animal experiments were performed in accordance with the United Kingdom Home Office Animals (Scientific Procedures) Act of 1986. *Alox15^-/-^* mice were bred under License (PC0174E40) at Cardiff. Weaned mice were housed in scantainers on a 12-hour light/dark cycle at 20 - 22 °C, with free access to normal chow and water. Mice were transferred to Bristol University for injury model carried out in line with United Kingdom Home Office approval in accordance with the Animals (Scientific Procedures) Act of 1986 (PPL30/3445). Sample sizes were based on previously published data, with n = 8 per group, and experiments were not blinded ^38^. Mice were anaesthetized (100 mg/kg ketamine and 10 mg/kg xylazine) and administered Dylight-488–anti-GPIbb (Emfret Analytics, 0.1 mg/g) into the jugular vein to label platelets. Carotid arteries were surgically exposed, and a piece of Whatman filter paper (2 × 1 mm) saturated with 24 % ferric chloride solution applied on the artery for 1 min. Thrombus formation was recorded for 10 min using fluorescence time-lapsed microscopy (Olympus BX51-WI microscope coupled with a Rolera XR digital camera). Data were acquired using Streampix 4 software (v4.24.0, Norpix, Montreal, Canada). Digital time-lapsed images were analysed using ImageJ (v1.53) to generate the integrated fluorescence density of each time frame as described previously^38, 39^.

### Lipid extraction, and liquid chromatography-tandem mass spectrometry (LC/MS/MS)

*(i) blood cells and EVs.* Internal standards (IS) were added (10 ng 1,2-dimyristoyl-PE and -PC, Avanti Polar Lipids) to leukocytes, platelets or EV samples, then lipids were extracted using a solvent mixture [1 M acetic acid, 2-propanol, hexane (2:20:30)] at a ratio of 2.5 ml of solvent to 1 ml sample, vortexing 1 min, then adding 2.5 ml of hexane. Following 1 min vortexing and centrifugation (400 g, 5 mins), lipids were recovered in the upper hexane layer. The samples were then re-extracted by the addition of hexane (2.5 ml/ml sample) followed by further vortexing and centrifugation, as above. The combined hexane layers were dried under vacuum and stored at -80 °C until analysis using LC/MS/MS.
*(ii) Clot lipid extraction.* Clots were retrieved onto dry ice and divided with a scalpel if deemed to be larger than 200 mg by visual estimate. Next, clots were weighed, transferred into 1.5 ml tubes and placed on wet ice. Since these are clots that contain high levels of pro-oxidant hemoglobin, ice cold anti-oxidant buffer (0.5 ml PBS containing 100 μM DTPA, 100 μM BHT and 7.5 μM acetaminophen), IS (10 ng DMPC/DMPE) and 8-10 ceramic beads (1.4 mm) were added to minimize artefactual oxidation that could take place during tissue processing. To reduce unstable lipid hydroperoxides, SnCl_2_ (100 mM) was added. The tubes were placed in a Bead Ruptor Elite^TM^ (Omni, USA) and tissue homogenization carried out at 4 m/s for 20 seconds at 4 °C. Homogenized clot samples (0.5 ml) were transferred to glass tubes containing 2.5 ml ice-cold methanol (MeOH) and placed on ice. A further 0.5 ml anti-oxidant buffer was added. A modified Bligh and Dyer lipid extraction method was carried out by adding 1.25 ml chloroform per sample. The mixture was vortexed for 1 min, and samples were placed on ice for 30 min. Following this, 1.25 ml of chloroform was added and the mixture vortexed for 1 min. 1.25 ml of H_2_O was added, solvent mixture vortexed for 1 min and then centrifuged at 400 g for 5 min at 4 °C to support phase separation. The bottom layer was collected into fresh vials using glass Pasteur pipettes and samples were evaporated to dryness using a Rapidvap vacuum evaporation system (Labconco, USA). Lipids were resuspended in 100 μl MeOH, transferred to HPLC vials, and stored at -80 °C until analysis using LC/MS/MS.

For LC/MS/MS, samples were separated on a C18 Luna, 3 μm, 150 mm x 2 mm column (Phenomenex) gradient of 50 - 100 % solvent B for 10 min followed by 30 min at 100 % B (Solvent A: MeOH:acetonitrile:water, 1 mM ammonium acetate, 60:20:20; Solvent B: MeOH, 1 mM ammonium acetate) with a flow rate of 200 μl/min. Products were analyzed in multiple reaction monitoring (MRM) mode, on a Q-Trap 6500 (Sciex) operating in negative mode (Supplementary Methods). The area under the curve for the analytes was integrated and normalized to the IS. For quantification of HETE-containing PL, standard curves were generated using PC and PE 18:0a/HETE, as described previously^40^. Limit of quantitation (LOQ) is defined as signal:noise of 5:1 with at least 6 data points across a peak. Settings were dwell time of 75 msec, declustering potential (DP) -50 volts, entrance potential (EP) -10 volts, collision energy (CE) -38 volts, and collision cell exit potential (CXP) -11 volts. Multiple reaction monitoring (MRM) transitions for 48 platelet eoxPL, were analysed for platelets from the health cohort^41^, while for the clinical samples and clots, only the most abundant HETE-PL were analysed. We note that for many of these lipids, due to the absence of full structural information and synthetic standards, their names are annotated only in part, e.g. by omitting the position of hydroxyl group or avoiding naming the specific oxygenated functional group where this information was not available, and they are calculated as area of analyte:internal standard (A/IS, cps). The list of MRM transitions used are provided in Supplementary Tables 4 (healthy cohort) and 5 (clinical study).

### LC/MS/MS of 12-HETE and TxB_2_

Platelet lipid extracts from the healthy cohort were analysed for 12-HETE and TxB_2_ using LC/MS/MS. However, since samples had already been extracted for eoxPL analysis, this was a post-hoc estimate, meaning that IS (5 ng of each of 12S-HETE-*d8* and TxB_2_-*d4* (Cayman Chemical, USA)) were added after lipid extraction. Thus, variability in extraction efficiency would not be accounted, and data are expressed as A/IS (area, cps). Also, insufficient volumes of all samples were available due to prior assays, and so only a subset of 14 were measured. Samples were processed on a C18 Spherisorb ODS2, 5 μm, 150 x 4.6-mm column (Waters, Hertfordshire, UK) using a gradient of 50 – 90 % B over 10 min (A, water:acetonitrile:acetic acid, 75:25:0.1; B, MeOH:acetonitrile:acetic acid, 60:40:0.1) with a flow rate of 1 ml/min. LC/MS/MS was by electrospray ionization on a Q-Trap 4000 (Sciex, UK) operating in negative ion mode, monitoring for MRM transitions for 12-HETE, TxB_2_, and their respective deuterated IS. Instrument settings are in Supplementary Table 6. Limit of quantitation (LOQ) is defined as signal:noise of 5:1 with at least 6 data points across a peak.

## Chiral LC/MS/MS of clot HETEs

Lipid extracts underwent alkaline hydrolysis followed by chiral LC/MS/MS as described previously^10, 42^. 80 µl of lipid extract was dried under N_2_, then resuspended in 1.5 ml 2-propanol and vortexed for 1 min. 1.5 ml of 1 M NaOH was added to followed by vortexing for 10 sec. Samples were placed in a 60 °C water bath for 30 min. 140 µl of 12 M HCl was added to acidify to pH 3.0. Next, 3 ng 12(S)-HETE-*d8* were added to each sample, followed by the addition of 3 ml hexane, vortexing for 1 min and centrifuging at 250 g for 5 min at 4 °C. Lipids were recovered in the upper hexane layer. The aqueous portion was re-extracted by addition of a further 3 ml hexane, vortexed for 1 min and centrifuged at 250 g for 5 min at 4 °C. The hexane layers were combined, and dried using a Rapidvap (Labconco Corporation, USA). Lipids were resuspended in 60 μl MeOH and transferred to an LC/MS/MS vial. The *S* and *R* enantiomers for HETE positional isomers were separated on a Chiralpak AD-RH, 5 μm, 150 x 4.6-mm column (Daicel, France) using an isocratic solvent (MeOH:H_2_O:glacial acetic acid, 95:5:0.1 v/v) with a flow rate of 0.3 ml/min over 25 min. Products were quantified by LC/MS/MS on an Sciex Q-Trap 4000 in negative ion mode using MRM transitions for 15-, 12-, 11-, 8- and 5-HETE, as well as 12(S)-HETE-*d8,* with instrument settings as shown (Supplementary Table 7)

## Statistical analysis

LC/MS/MS data was exported from Analyst v1.6.3 (Sciex, USA) and analysed using MultiQuant v3.0.3 (Sciex, USA). Statistics and bioinformatics used the coding environment R (Open source, version 3.5.3). For heatmaps, mean values (ng/ml or cell number) were log10 transformed and plotted as intensity values using the Pheatmap package with lipid hierarchical clustering. Each lipid was annotated, using a color code, according to its presumed enzymatic origin^43^. For box plots, edges indicate the interquartile range (IQR) with the median line inside the box. Whiskers indicate 1.5 times the IQR.

For the clinical samples, significance was determined using Mann-Whitney-Wilcoxon Test for pairwise comparison and Kruskal-Wallis test for analysis of differences between more than two groups. Tests were done with base R. Where determined to be normally distributed using Kolmogorov-Smirnov test (https://www.socscistatistics.com/tests/kolmogorov/default.aspx),

ANOVA with Tukey post hoc (https://astatsa.com/OneWay_Anova_with_TukeyHSD/_get_data/) or Students t-test were used as stated in legends. For the healthy control cohort, data were normally distributed, so Students t-test was used for paired samples. Significance was designated p-value <0.05. Lipids which were undetected in more than 80% of (activated) samples were excluded from the analysis. For the purposes of statistical comparison, zeros were replaced with 50% of the LOQ value as calculated by a standard curve on the same instrument and column settings. Multiple comparison adjustment was not used since variable numbers were <50. Principal component analysis (PCA) used the singular value decomposition method provided by the base R ‘prcomp’ function. Example chromatograms for all lipids detected are provided in Supplementary Figure 1. Data reporting is in line with ARRIVE^44^ and STROBE^45^ guidelines.

## Results

Platelets from all patient groups generate higher levels of 12-HETE-PL but reduced levels of 11- and 15-HETE-PLs, than healthy controls.

HETE-PL were quantified in washed platelets from patients with ACS, CAD and RF basally and following activation with thrombin. We focused on the most abundant PE and PC species previously detected in platelets; four PEs and two PCs which contain 18:0, 18:1 or 16:0, including both plasmalogen and acyl forms, while scanning for five HETE positional isoforms (5-, 8-, 11-, 12-, 15-) ^10^. Little or no HETE-PL were detected basally, however thrombin stimulated large increases, particularly for abundant 12-HETE-PL species formed via the action of 12-LOX^10^ (Figure 1 A). Other HETE-PLs also increased on activation, mainly 15- and 11-HETE-PL but also very low levels of 5- and 8-HETE-PL (Figure 1 B,C, Supplementary Figure 2 A,B). Amounts of 12-HETE-PL generated were significantly higher in activated platelets from all patient groups, than HC (Figure 1A). The elevation in 12-HETE-PL was driven exclusively by higher levels of the three diacyl-12-HETE-PL, with no significant differences for the plasmalogens (Figure 1 D-F, Supplementary Figure 2 C-E). In contrast, 15- and 11-HETE-PLs were significantly reduced in activated platelets from all groups compared to HC (Figure 1 B,C). Here, both plasmalogen and acyl-forms of HETE-PEs, and PCs were impacted equally (Supplementary Figures 2 F-K, 3 A-F). 5- and 8-HETE-PL in activated platelets were not different for patient groups versus healthy controls (Supplementary Figure 2 A,B). Overall, this indicates that diacyl-eoxPL from 12-LOX are significantly elevated in patient groups, while both acyl and plasmalogen 15- and 11-HETE-PL species are reduced. The enzymatic origin of these lipids will be explored below.

**Figure 1:**
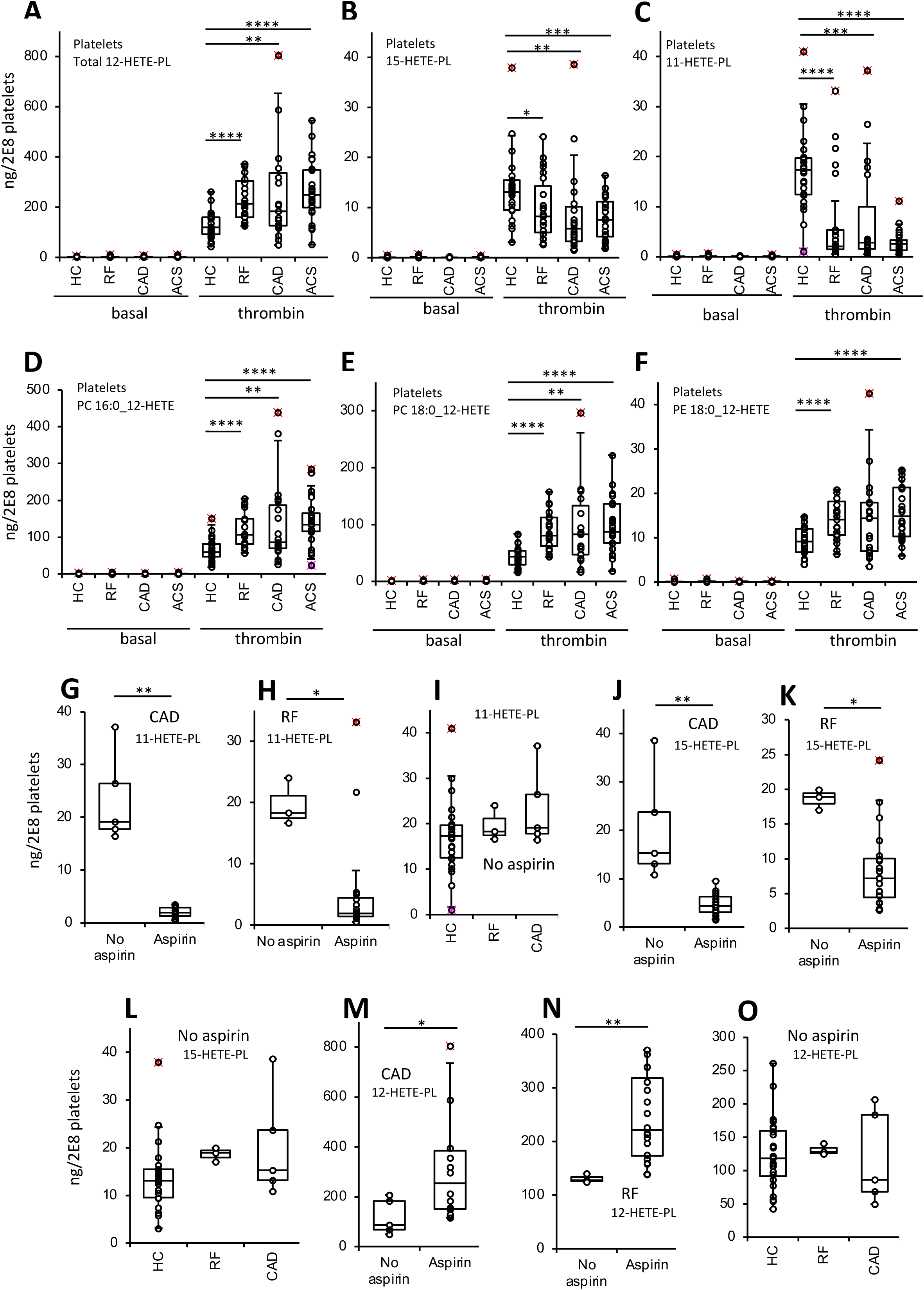
Aspirin reduces COX-1 generated eoxPL but increases 12-LOX generated acyl eoxPL in thrombin-activated platelets in arterial thrombosis. Blood samples from a clinical cohort of arterial thrombosis were collected, platelets (2 x 10^8^/ml) isolated, and then thrombin-activated (0.2 unit·ml^−1^ thrombin, 37 °C, 30 min). Lipids were extracted and analysed using LC/MS/MS and HETE-PLs quantified. Individual isomers were quantified then totaled to give amounts as follows: *Panel A*. 12-HETE-PL, *Panel B*.11-HETE-PL, and *Panel C.*15-HETE-PL. ACS: acute coronary syndrome (n = 24), CAD: coronary artery disease but no ACS (n = 19), RF: Risk factors with no significant coronary artery disease (n = 23), HC: Healthy control (n = 24). *Panels D-F.* Lipids are displayed as individual diacyl-12-HETE-PL. *Panels G,H.* 11-HETE-PL were compared across RF and CAD groups based on whether they were on aspirin (n = 5, 14, Panel G, n = 3, 20, Panel H for no aspirin and aspirin respectively). Panel I. 11-HETE-PL were compared in patients not on aspirin, in HC, RF and CAD groups (n = 23, 3, 5, for HC, RF and CAD respectively). *Panels J, K.* 15-HETE-PL were compared across RF and CAD groups based on whether they were on aspirin (n = 5, 14, Panel J, n = 3, 20, Panel K for no aspirin and aspirin respectively). *Panel L*. 15-HETE-PL were compared in patients not on aspirin, in HC, RF and CAD groups (n = 23, 3, 5, for HC, RF and CAD respectively). *Panels M,N.* 12-HETE-PL were compared across RF and CAD groups based on whether they were on aspirin (n = 5, 14, Panel M, n = 3, 20, Panel N for no aspirin and aspirin respectively). *Panel O*. 12-HETE-PL were compared in patients not on aspirin, in HC, RF and CAD groups (n = 23, 3, 5, for HC, RF and CAD respectively). Statistical significance was tested with Mann-Whitney-Wilcoxon test for pairwise comparison or Kruskal-Wallis test for non-parametric analysis of variance on rank (* p <0.05, ** p <0.01, *** p <0.001).

**Figure 2:**
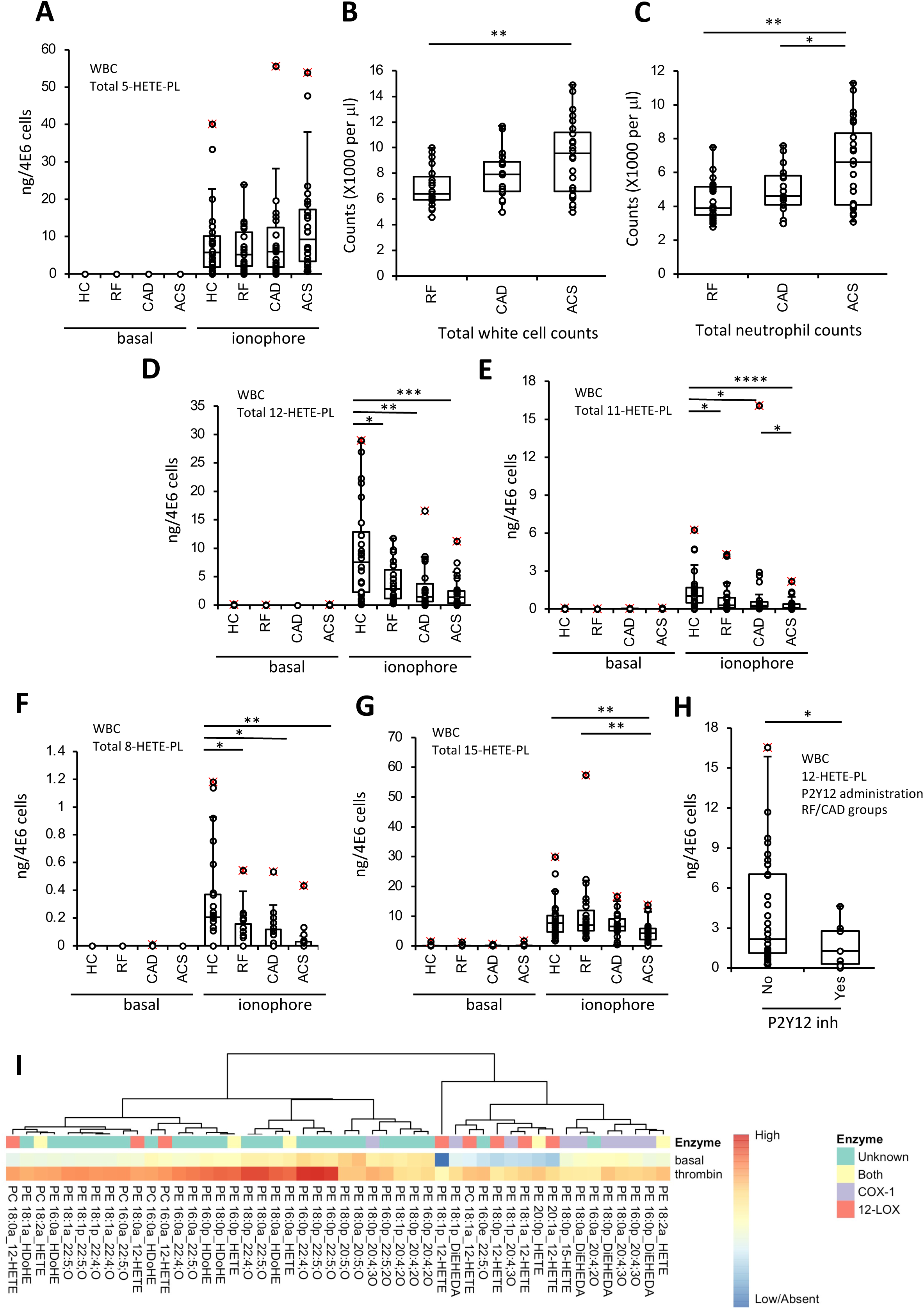
12-,11- and 15-HETE-PL in ionophore-activated leukocytes are lower in patients with arterial thrombosis than healthy controls, and thrombin stimulates generation of many eoxPL in healthy subjects. Blood samples from a clinical cohort of arterial thrombosis were collected, leukocytes (4 x 10^6^/ml) isolated, and then ionophore-activated (10 μM A23187, 37 °C, 30 min). ACS: acute coronary syndrome (n = 24), CAD: coronary artery disease but no ACS (n = 19), RF: Risk factors with no significant coronary artery disease (n = 23), HC: Healthy control (n = 24). Lipids were extracted and analysed using LC/MS/MS and HETE-PLs quantified. Individual isomers were quantified then totaled to give amounts as follows *Panel A*: 5-HETE-PL. *Panels B,C.* Leukocyte and neutrophil counts were obtained from clinical laboratory measurements. *Panel D:* 12-HETE-PL, *Panel E*: 11-HETE-PL, *Panel F:* 8-HETE-PL, *Panel G*: 15-HETE-PL. *Panel H:* 12-HETE-PL levels in RF and CAD groups compared with/without P2Y12 inhibitor supplementation (n = 33, 9 respectively). Statistical significance was tested with Mann-Whitney-Wilcoxon test for pairwise comparison (*: p <0.05, **: p <0.01, ***:p <0.001). *Panel I. Heatmap showing generation of diverse eoxPL in healthy subjects.* Blood was collected from 28 healthy volunteers (14 male, 14 female) following an NSAID washout period, and platelets isolated and activated *in vitro* using thrombin, as outlined in Methods. Levels of 48 (most abundant) eoxPL were profiled using LC/MS/MS, with analyte:internal standard (A/IS, cps) determined. Log10 values were calculated and plotted using Pheatmap in R as outlined in methods, with assignment to enzymatic sourced as outlined.

**Figure 3:**
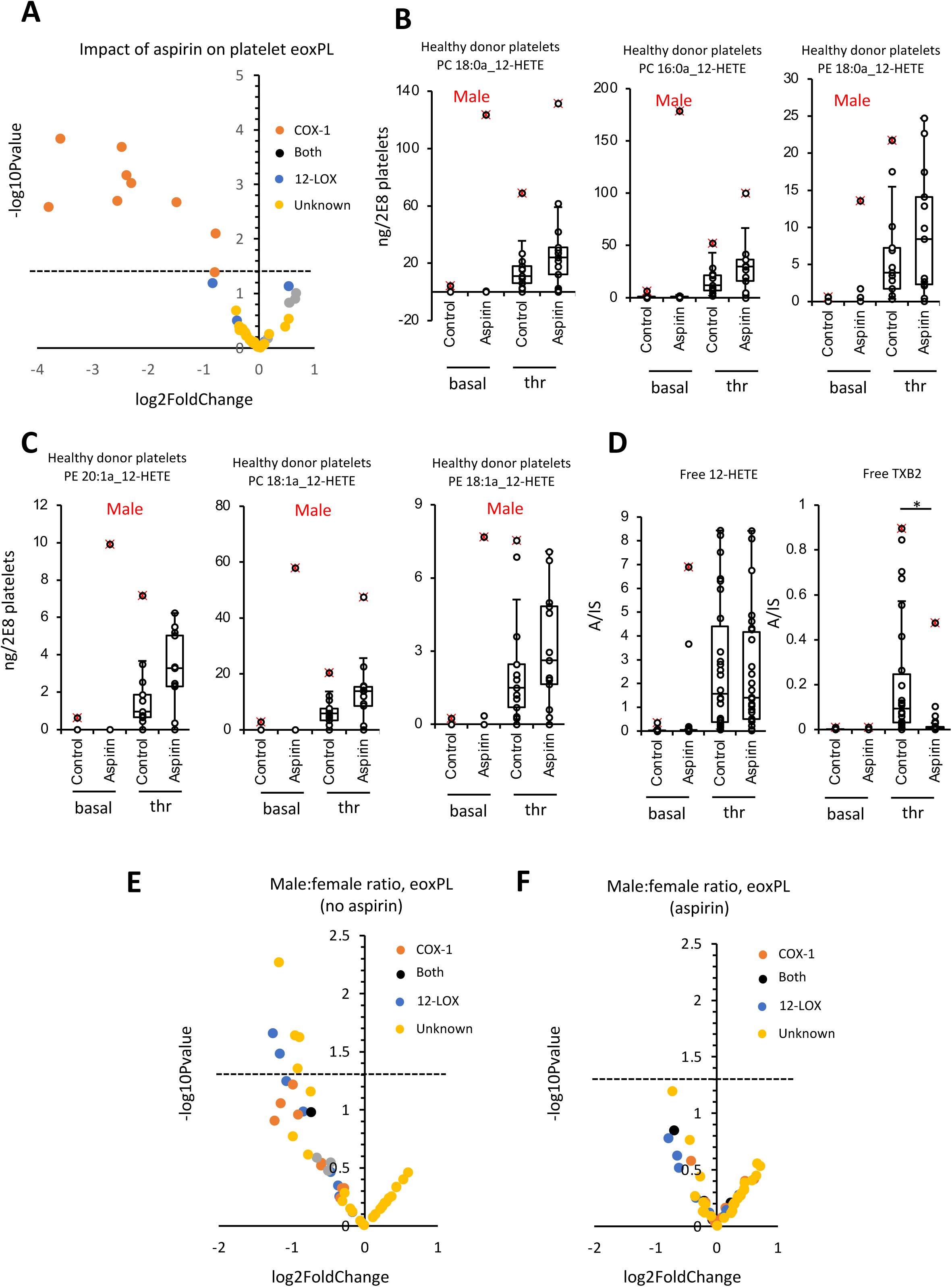
Aspirin supplementation reduces COX-derived eoxPL, and increases diacyl 12-HETE-PL from LOX, in platelets from a healthy cohort. Blood was collected from 28 healthy volunteers (14 male, 14 female) following an NSAID washout period, and platelets isolated and activated *in vitro* using thrombin, as outlined in Methods. Subjects next were administered aspirin (75mg/day) for a week, before repeat sampling of platelets. Lipids were extracted from either resting or thrombin-activated platelets, and analysed using LC/MS/MS, with analyte:internal standard (A/IS, cps) determined as outlined in Methods. *Panel A. Volcano plot showing impact of aspirin on platelet eoxPL.* Data from the healthy cohort eoxPL was analysed to generate log2fold change and -log10Pvalues, which were plotted with lipids labelled according to their enzymatic origin. *Panels B, C. Box plots showing diacyl 12-HETE-PL molecular species in platelets from males are elevated by aspirin supplementation*. Lipids were extracted and analysed using LC/MS/MS as outlined in Methods. *Panels D,E. Free 12-HETE and TxB2*. These were measured in platelet lipid extracts using LC/MS/MS (n = 14, 50:50 male female) as outlined in Methods. *Panels D,E. Platelets from males generate lower levels of several eoxPL than females*, *in an aspirin-sensitive manner.* Lipids were plotted on a volcano plot as -log10Pvalue and log2FoldChange, male:female, labelled by pathway of origin. Statistical significance was tested with T-test for pairwise comparison and ANOVA for within-group differences (*: p <0.05, **: p <0.01, ***:p <0.001, ns or no stars: not significant).

Aspirin supplementation associates with elevated 12- and reduced 11- and 15-HETE-PLs in platelets from patient groups.

In platelets, 15- and 11-HETE-PL likely arise from COX-1 formation of 15- and 11-HETE, followed by their esterification into PL. To test this, the impact of aspirin supplementation on their formation was determined. First, we noted that all ACS, but no HC participants were taking aspirin (Supplementary Table 8). In contrast, in both the RF and CAD groups, a small number were not on aspirin allowing a direct comparison within these groups. Here, a significant suppressive effect of aspirin on both 11- and 15-HETE-PLs was observed, while for participants not on aspirin in the HC, RF and CAD group, there were no significant differences (Figure 1 G-L). Aspirin treatment was also associated with a significant increase in 12-HETE-PLs in the RF and CAD groups (Figure 1 M,N), while for participants not on aspirin, there was no impact of disease (Figure 1 O). Next, the individual isoforms for 12-HETE-PLs in the RF and CAD groups were measured. In the case of 12-HETE-PLs, two of the three diacyls forms were significantly elevated by aspirin supplementation, while for all plasmalogens, there was no effect (Supplementary Figure 3 G,H). All female RF and CAD patients were on aspirin, thus, to control for gender, males only were compared. As above, aspirin significantly enhanced platelet generation of diacyl 12-HETE-PLs for both RF and CAD groups combined (Supplementary Figure 4).

In contrast, reductions in 11- and 15-HETE-PLs were seen for both acyl and plasmalogen forms in CAD and RF groups (Supplementary Figure 5). This strongly suggests that altered platelet eoxPL generation is at least in part related to aspirin. This idea will be later tested directly using a healthy cohort. Using clinical data, eoxPL generation by patient platelets was also compared to either medication use (anti-coagulants, P2Y12 inhibitors, statins) or other disease (diabetes, hypertension), but no correlations were noted.

### eoxPL generation was modulated by vascular disease in ionophore-activated leukocytes

Next, HETE-PL levels were analysed in total leukocyte populations isolated from patient groups and healthy controls, basally and following ionophore activation to determine the capacity of the cells to activate maximally. Here, HETE-PL were generally undetectable basally, but several formed on activation, mainly 15-, 12- or 5-HETEs, but also 8- or 11-HETE-PLs (Figure 2, Supplementary Figures 6-8 A). Overall, total 5-HETE-PL (when normalized to leukocyte number) generated by 5-LOX showed a small non-significant trend to elevate in all groups (Figure 2 A, Supplementary Figure 6 A). When cell counts for the patient groups were compared, a significant increase was seen for ACS versus RF for both total white cells and neutrophils (Figure 2 B,C). Thus in vivo, the amounts of 5-HETE-PE generated overall would be further elevated in line with the higher cell counts. Several 15, 12, 11, or 8-HETE-PL species were significantly decreased in patient groups, versus HC (Figure 2 D-G, Supplementary Figures 6 B, 7, 8 A). We next examined if these reductions were associated with drug treatments or co-existing conditions, however there were no significant differences seen for aspirin, statins, diabetes or hypertension (Supplementary Figure 8 B). The exception was 12-HETE-PL which was significantly reduced in white cells from RF/CAD patients on P2Y12 inhibitors (Figure 2 H). This observation is likely related to leukocyte:platelet aggregates present *in vivo*, as discussed later.

### Characterizing the impact of aspirin and gender on eoxPL generation by human platelets from healthy volunteers

To further test the impact of aspirin, we next turned to a healthy cohort. Previously, in platelets from three unrelated healthy human donors, ∼100 eoxPL were seen to be generated acutely on thrombin activation^41^. Some of these were already characterized eoxPL, but many remain incompletely annotated with only partial structural information on the oxylipin, specifying the chain length, number of rings/double bonds, and oxygenation, e.g. 22:4,2O^41^. Taking this further, we expanded the study up to a cohort of 28 subjects (50:50 male:female), testing the impact on eoxPL of (i) aspirin treatment, (ii) gender and (iii) repeated sampling at 2 monthly intervals. Subjects first provided a baseline blood sample following an NSAID washout period, and then immediately following one week on 75mg/day aspirin, and the generation of the 48 most abundant eoxPL previously shown to be generated by thrombin-activated platelets were measured (Supplementary Table 9)^41^. Lipids were putatively assigned to be from either COX-1 or 12-LOX based on known sources of the oxylipins in platelets, e.g. 12-HETE (12-LOX), or 11-, 15-HETE, DiEHEDA (COX-1). We also assigned oxylipins containing 20:4;3O as being likely prostaglandins, e.g. PGE_2_, which are known to be generated in eoxPL forms by platelets^46^. Across the panel, 45 out of 48 lipids were significantly elevated on thrombin activation (Figure 2I). Next, to analyze the impact of aspirin supplementation on thrombin stimulation of eoxPL generation, platelets were obtained before and after 1 week on 75 mg/day aspirin in the same individuals. The lipids proposed to originate from COX-1 were all significantly suppressed (Figure 3 A, orange markers). This data confirms the enzymatic origin of putative COX-1 derived eoxPL. 12-HETE containing eoxPL increased following aspirin supplementation, although this didn’t reach significance (Figure 3 A, blue markers). When analysed for gender differences, the median diacyl HETE-PLs were consistently elevated in males following aspirin treatment, although this didn’t reach significance (Figure 3 B,C). In contrast, aspirin had no discernible effect on diacy-12-HETE-PL generation in females, or in plasmalogen 12-HETE-PLs in either gender (Supplementary Figure 9 A). This small increase in diacyl-12-HETE-PLs is similar to that seen in the clinical cohort, further supporting the idea that it relates to aspirin. The small elevations of diacyl-12-HETE-PL could result from either elevated generation of the free 12-HETE precursor or increased rates of esterification of 12-HETE into lysoPL, by ACSL/LPLAT enzymes^47^. To test this, levels of 12-HETE and TXB_2_ were analysed, however while TXB_2_ generation was suppressed around 95% by aspirin, there was no elevation in free 12-HETE for either males or females (Figure 3 D, Supplementary Figure 9 B). This suggests that esterification of 12-HETE to form diacyl, but not plasmalogen-HETE-PLs is slightly enhanced by aspirin, in line with our observations in the cardiovascular cohort shown earlier.

Next, the influence of gender (in the absence of aspirin) was tested, and showed that many lipids were overall lower in platelets from males, with six significantly reduced (Figure 3 E, lipids with -log10Pvalue >1.3). These were either from LOX or unknown pathways, although all COX-1 or LOX derived eoxPL tended to be somewhat reduced indicating a lower overall thrombin response in males. The significantly-reduced species were two diacyl 12-HETE-PLs (from LOX, PC 18:1a_12-HETE, PC 16:0a_12-HETE) with three others being plasmalogen-PEs containing 20:4;2O (PE 18:0p_20:4;2O, PE 16:0p_20:4;2O, PE 18:1p_20:4;2O). These may represent diHETEs, from CYP/sEH. None of these were directly impacted by aspirin in either males or females, indicating they are not generated by COX-1 directly. However, when subjects were on aspirin, this gender effect was attenuated and male and female generation of eoxPL became rather similar (Figure 3 F). Gender differences in platelet reactivity are well described in the literature, and will be summarize in the Discussion.

### EoxPL from COX and LOX show seasonal variations which are abrogated by aspirin

The inter-individual variability of eoxPL, when repeatedly sampled over time, is unknown. To test this, a subgroup of healthy volunteers (7 male, 7 female) returned at two subsequent two monthly intervals to provide additional samples pre and post aspirin. When subjects were not on aspirin, platelet generation of eoxPL from COX/LOX were up to 2.5 - 3-fold higher in spring versus autumn, a relatively modest fluctuation (Figure 4 A). In contrast, most lipids assigned as “unknown” did not show any seasonal variation (Figure 4 A). When subjects were on aspirin, the increase in COX-derived lipids in spring was not seen (Figure 4 B), although for LOX-derived eoxPL the seasonal trend remained, and as before “unknowns” were unaffected.

**Figure 4:**
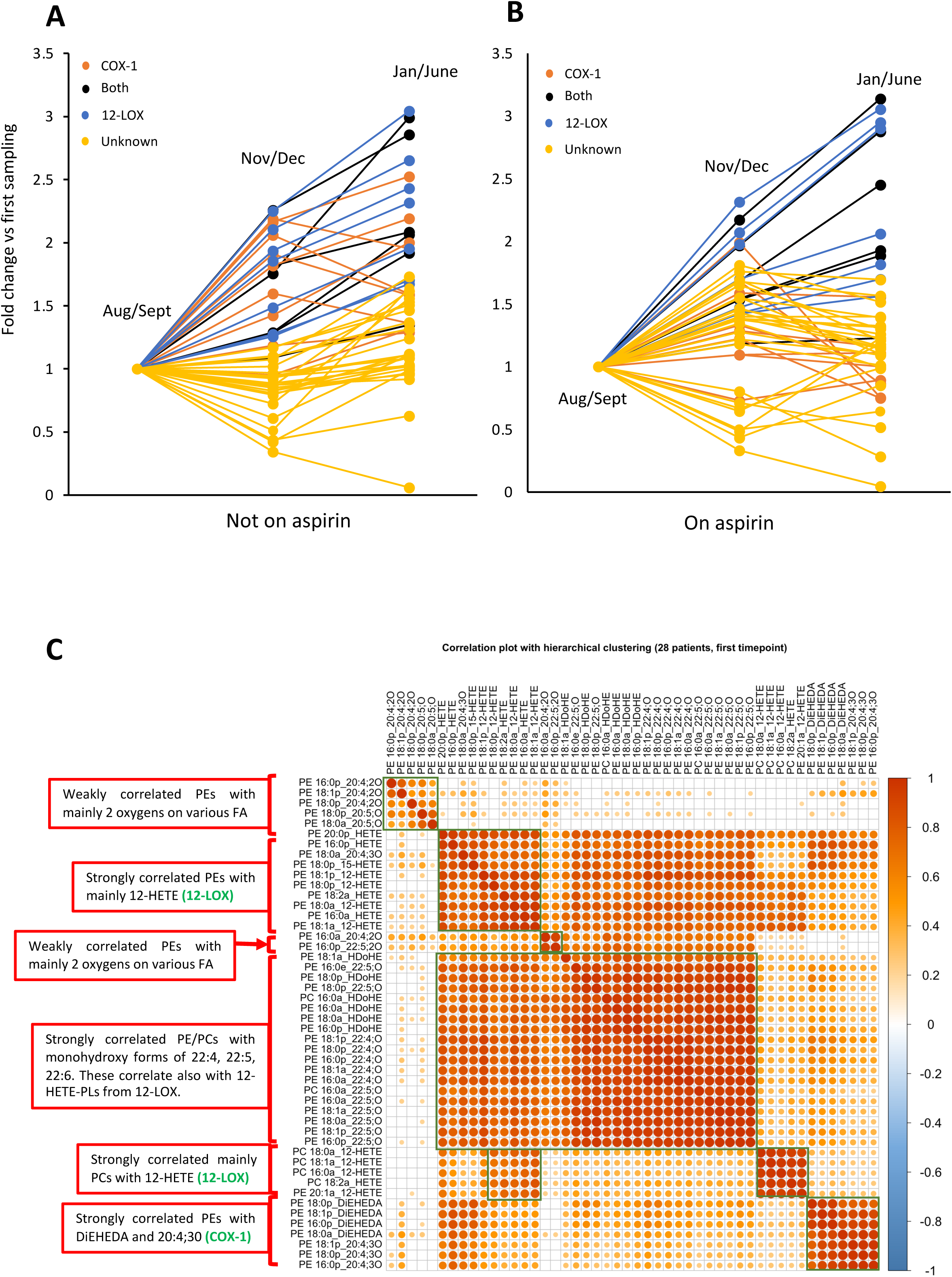
EoxPL generated by COX show seasonal variation that is attenuated by aspirin supplementation. Healthy volunteers (n = 14) donated platelets before and after 1 week aspirin (75mg/day) supplementation. The donation protocol was repeated after 2-3 months, to generate repeat samples for each volunteer. Lipids were extracted from platelets (2 x 10^8^) basally or following thrombin activation (0.2 U/ml) as in Methods. Extracts were analysed using LC/MS/MS, calculating A/IS values for each lipid at each timepoint, with volunteers either off (Panel A) or on (Panel B) aspirin. *Panel C. Pearson’s correlation coefficients demonstrate clustering of eoxPL by Sn2 fatty acyl composition and headgroups.* Pearson’s correlations were calculated, and significant (p<0.05) correlations plotted.

### EoxPL correlate based on Sn-2 oxylipin composition

To assess biochemical relatedness of eoxPL molecular species, correlation plots were generated for eoxPL using the data from the first sampling time point (n = 28, basal, thrombin, +/- aspirin supplementation). Here, groupings aligned with *Sn*-2 fatty acyl oxylipin structure strongly (Figure 4 C). For example, PEs containing HETEs, HDOHEs or other monohydroxy lipids clustered together in individual groups. PC with 12-HETE formed a separate cluster, indicating influence of the headgroup on biochemical behavior. Lipids from COX-1 correlated closely but were very distinct from other lipids, most likely due to the selective impact of aspirin. Last, two groups which contained eoxPL with 2 oxygens were seen. These groups weakly clustered together, but were very distinct from others, suggesting a different enzymatic source, potentially CYP450/sEH. There were no negative correlations since all lipids either correlated positively or not at all.

### Lipidomics demonstrates higher levels of platelet eoxPL in human arterial limb and coronary (STEMI), than carotid thrombi

Next, to study in vivo generated eoxPL, arterial thrombi removed from patients were analysed, comparing three anatomical sites: carotid, limb and STEMI. Here, clots surgically obtained from patients were homogenized, lipids extracted and HETE-PL measured using LC/MS/MS, followed by hydrolysis of fatty acyls and analysis of HETEs for chirality to confirm enzymatic origin. 12-HETE from platelet 12-LOX, 15-HETE from leukocyte 15-LOX and 5-HETE from neutrophil 5-LOX should be primarily *S*, while 15-HETEs from COX-1 can be either *R* or *S*^48, 49^. 11-HETE generated by COX-1 is predominantly *R*^48, 50^.

Although from a small group of patients with baseline differences due to age, demographics, and clinical background, there were clear signatures related to anatomical site as follows. In carotid clots, 15-, 11- and 5-HETE-PL were the prominent eoxPL with lower levels of 12- and 8-HETEs detected. This signature suggested white cells as the prominent source of eoxPL (Figure 5 A). Chiral analysis of carotid clot lipids showed that 12-, 15- and 5-HETEs were all slightly more prominent as *S* isomer forms (55-60 %) while both 8- and 11-HETEs were around 50 % *S/R*. Coupled with the higher levels of 15-HETE-PL *versus* 12- or 8-HETE-PLs, we suggest that 15-HETE-PL originates from both leukocyte 15-LOX (*S*) and COX (*S/R*), while some of the 5-HETE-PL is from neutrophil 5-LOX (*S*) (Figure 5 B). The other HETE-PLs;12-, 8- and 11- may be non-enzymatic derived. There was notable absence of 12-HETE-PL in carotid clots, suggesting little or no platelet involvement (Figure 5 A). In contrast, STEMI and limb clots were characterized by higher levels of 12-, followed by 15- and 11-HETE-PLs, relative to lower amounts of 5- and 8-HETE-PLs. This indicated a platelet signature, largely dominated by 12-LOX (12-HETE-PL) with some COX-1 derived eoxPL (11-, 15-HETE-PL). As confirmation, limb and STEMI clots contained around 95 % 12*S*-HETE (Figure 5 B). In limb clots, 11-HETE was around 75 % the *R* isomer, consistent with COX-1, although in STEMI, it was around 50 % (Figure 5 B). 8-HETE in limb and STEMI clots was around 75 % or 60 % *R* (Figure 5 B). Although non-enzymatic oxidation would predict 50 % S/R for 8-HETE, a similar pre-dominance of the *R* enantiomer in serum from clotted blood was previously shown^50, 51^, and 8R-HETE is a known side product of platelet 12-LOX^52^ . In limb and STEMI clots, 5-HETE-PL were relatively low in comparison to other positional isomers, and were around 50-60 % *S* isomer (Figure 5 B). Thus, they may be primarily non-enzymatic, with a small involvement of neutrophil 5-LOX. Notably, clots contain large amounts of hemoglobin which could directly contribute to non-enzymatic oxidation, either in vivo or during lipid extraction, despite inclusion of antioxidants and metal chelators during tissue processing.

**Figure 5:**
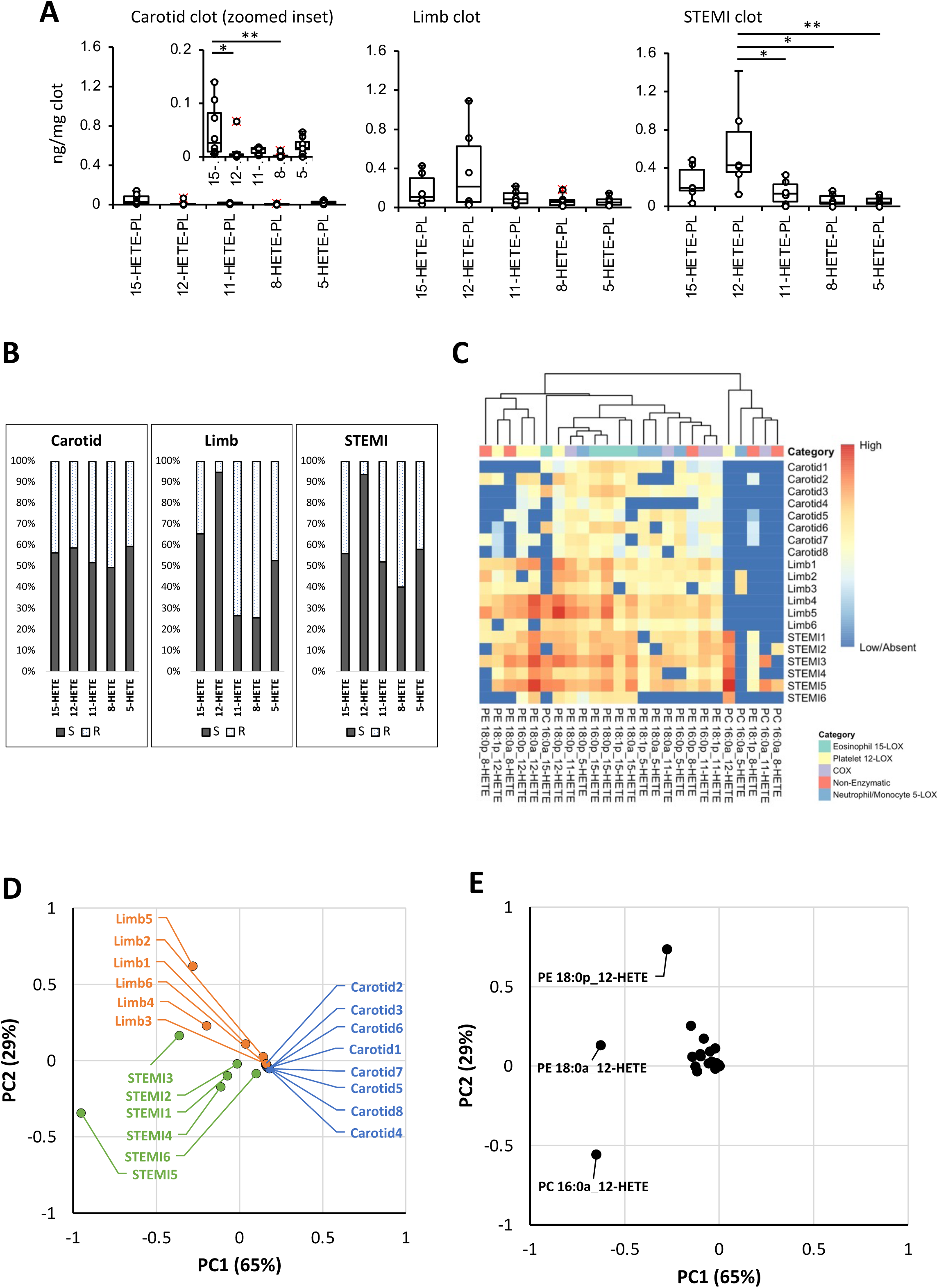
EoxPL profiles of limb and STEMI clots show a strong platelet signature while carotid clots are more abundant in white cell isoforms. *Panel A: The profile of arterial thrombi eoxPL shows site specific signatures.* Arterial thrombi were extracted, snap frozen on dry ice and frozen at -80 °C from patients undergoing angioplasty for ST-elevation myocardial infarction (STEMI, n = 6), carotid endarterectomy (carotid n = 8) or peripheral vascular embolectomy (limb n = 6). Clots were homogenized and lipids extracted as in Methods, then analysed using LC/MS/MS, quantified and normalized by tissue weight (ng lipid/100 mg clot). *Panel B: Chiral analysis confirms the platelet signature of limb and STEMI clots based on high S/R ratio of 12-HETE*. Clots (n = 3 each) lipid extracts were hydrolyzed, then analysed using chiral LC/MS/MS as in Methods. *Panel C: The eoxPL dataset plotted as a heatmap shows site specific features.* A heatmap with hierarchical clustering was plotted using the pheatmap R package after log10 normalization of data. *Panel D: Principal component analysis of arterial thrombi by HETE-PL profile shows clustering of clots with common origin.* eoxPL data was analysed using the prcomp function in R (v version 3.5.3) *Panel E. The corresponding loading plot shows the variables most influential in explaining the difference between samples (each dot represents a lipid)*.

**Figure 6:**
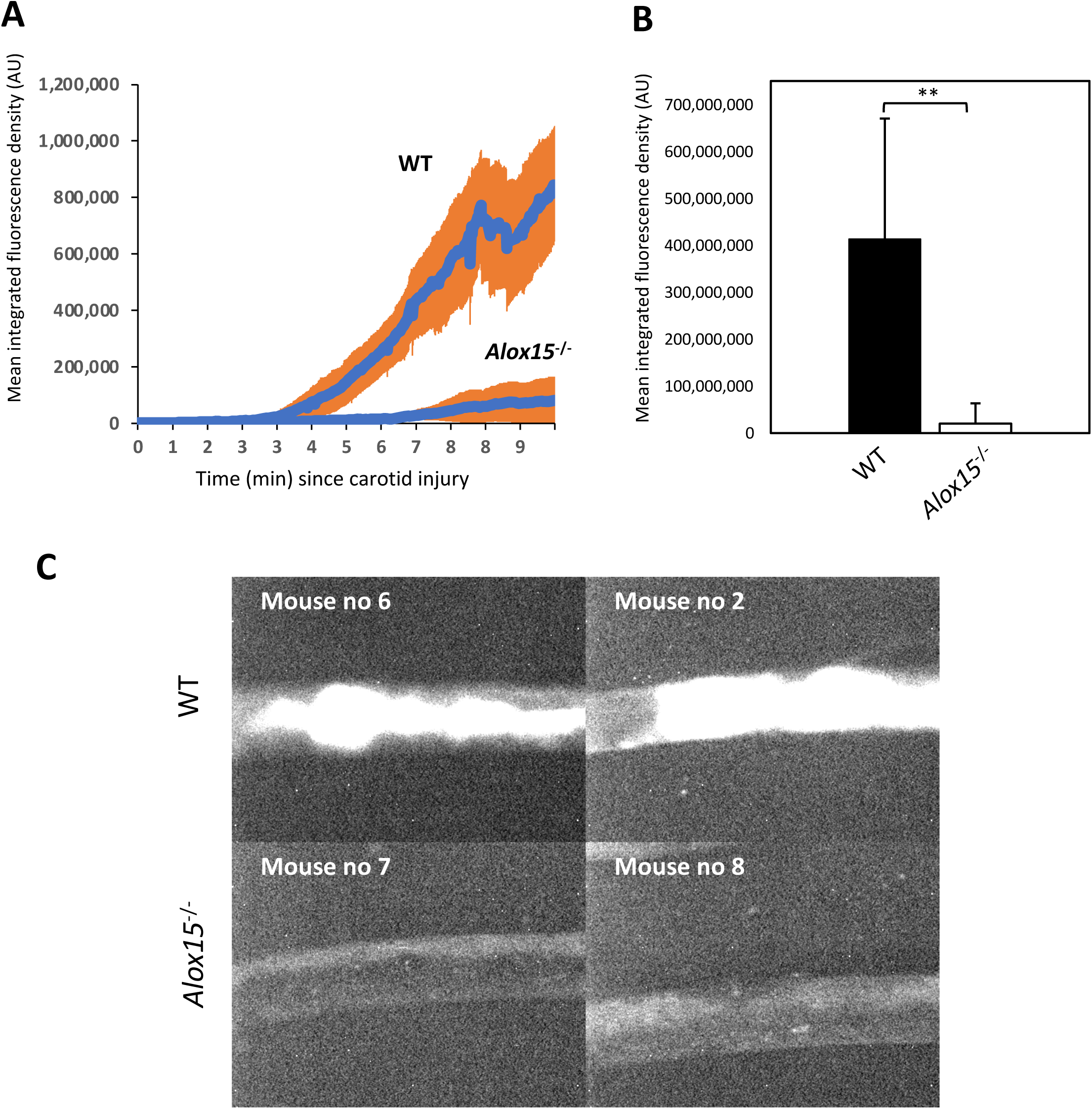
Alox15^-/-^ mice generate smaller carotid thrombi in an in vivo model. Panel A. Trace showing time course of thrombus formation, as mean integrated fluorescence (± *standard deviation), in Alox15^-/-^ mice compared with WT.* Female mice (12-week old) were anaesthetized and platelets labelled using Dylight-488–conjugated anti-GPIbb antibody. Carotid arteries were surgically exposed, and 24 % ferric chloride solution applied for 1 min. Thrombus formation was recorded for 10 min with fluorescence time-lapsed microscopy as described in Methods (n = 8 for each strain). Panel B. Integrated values for individual mice were calculated, then averaged. Statistical significance was tested with a 2-tail Student t-test. **p = 0.0079. *Panel C. Representative fluorescence images at the end of 10 min recording in two Alox15^-/-^ mice compared with 2 WT*.

The full dataset for all lipids is shown as a heatmap (Figure 5 C). Here, the higher abundance of 12- and 15-HETE-PL in STEMI and limb clots is visible, and the contribution of individual lipids can be seen. Next, a principal component analysis (PCA) was undertaken, to confirm which lipids contributed most strongly to the phenotype of the different clot types. This demonstrated separation of the STEMI and limb embolectomy samples in the PC1 direction (responsible for 65 % of the variance) (Figure 5 D). The loadings plot demonstrates that the HETE-PL lipids most responsible for this separation in PC1 between samples are three lipids containing the 12-HETE, namely two diacyl and one plasmalogen (Figure 5 E).

### Arterial thrombosis is significantly reduced in mice lacking Alox15

Herein, 12- and 15-HETE-PL were the predominant eoxPL in carotid or STEMI/limb clots, respectively and their levels varied in platelets from patients with arterial vascular disease. Previously we found that mice lacking ability to make these lipids due to genetic deletion of either *Alox12* or *Alox15* form smaller thrombi in venous models, however arterial thrombosis was not yet examined in vivo^8, 14^. Here, a ferric chloride carotid artery injury model was tested in mice lacking *Alox15*. This gene encodes 12/15-LOX in mice, which unlike its human orthologue (15-LOX), can generate 12- as well as 15-HETE-PL in murine leukocytes.

Following carotid injury, arterial thrombi formed at a significantly slower rate in *Alox15*^-/-^ mice compared with wildtype C57BL6 controls (Figure 6 A). The final thrombus size was also significantly smaller in mice lacking *Alox15* (Figure 6 B,C). These findings show that *Alox15*, potentially through forming 12- and 15-HETE-PL, significantly contributes to formation of arterial thrombi *in vivo*.

## Discussion

The regulation of pro-coagulant membranes of circulating blood cells in cardiovascular disease is poorly understood, both in relation to how they contribute to thrombotic risk, but also their involvement in formation of human clots *in situ*. Enzymatically oxidized phospholipids (eoxPL) were shown previously to be elevated in blood cells from patients with venous thrombosis, and be procoagulant *in vitro* and *in vivo*^53^. Up to now, their formation and potential role(s) in arterial thrombosis has not been investigated. Herein, using lipidomics, we characterize eoxPL in human arterial vascular disease in circulating blood cells and retrieved clots, as well as in a healthy cohort and demonstrate the importance of their biosynthetic pathway for mouse arterial thrombosis in vivo. Complex impacts of common anti-thrombotic/coagulant therapies on eoxPL generation were uncovered that can be summarized as: (i) inhibition of COX-1 derived eoxPL generation by aspirin, (ii) elevation of Lands cycle-dependent diacyl-12-HETE-PL generation in platelets by aspirin, (iii) reduction of platelet-derived eoxPL in leukocyte isolates by P2Y12 inhibitors, (iv) abrogation of gender and seasonal impacts on eoxPL generation in healthy volunteers by aspirin. These data show that aspirin’s impact on bleeding/thrombosis may move far beyond the traditional realm of platelet aggregation blockade, into modulation of membrane-dependent coagulation. How eoxPL modulation by aspirin/P2Y12 blockade ultimately impacts thrombotic risk via coagulation in humans in health and disease warrants further study.

In a human cohort of patients, thrombin activated platelet derived eoxPL generation showed complex changes that primarily associated with aspirin supplementation, findings which were then directly confirmed in a healthy cohort. While inhibition of COX-derived eoxPL will be due to direct inhibition of the enzyme, mechanistic reasons for elevations in LOX-derived species are not known. Here, the elevated LOX-derived eoxPL when taking aspirin could be due to a greater availability of arachidonate substrate as COX-1 is inhibited^54^. However, free 12-HETE generation by platelets was not elevated by aspirin in the healthy cohort, and furthermore only diacyl species were elevated. This indicates instead that their generation is upregulated at the level of esterification into PL via Lands cycle enzymes. This novel effect of aspirin requires further study, since the specific isoforms of ACSL/LPLATs that mediate 12-HETE acylation into diacyl-PL remain to be determined.

Due to the quantitative dominance of 12-HETE-PLs, the overall impact of aspirin was higher total levels of platelet pro-coagulant eoxPL detected. Importantly, resting platelets contain little or no eoxPL, with these mainly detected following agonist challenge (e.g. using thrombin). Thus, *in vivo*, while a patient is otherwise healthy, eoxPL lipids should be very low, unless a challenge occurs, e.g. through trauma, infection or plaque rupture. Where 12-LOX derived lipids are elevated by aspirin, they could potentially antagonize the anti-thrombotic impact of TXA_2_ inhibition, through promoting the pro-coagulant impact of the platelet membrane. The clinical impact of aspirin has always been considered to be primarily via its antiplatelet effects, not via modulation of coagulation per se. In support, washed platelets from patients on aspirin therapy still mount a pro-coagulant phenotype in vitro^55–57^. However, this does not exclude that eoxPL may influence coagulation under the conditions under which they are generated within the complex milieu of a thrombus in vivo. In line with this, a strong signature of platelet derived eoxPL was seen in retrieved human STEMI and limb clots, although it is not known at what stage of thrombus formation they were generated (e.g. early or late). In direct support of a role of these lipids in driving arterial coagulation in vivo, *Alox15^-/-^* mice, which lack many eoxPL, generated very small thrombi in an arterial model^15^, and higher levels of eoxPL are associated with pro-thrombotic autoimmunity^8^. Furthermore, we previously showed that both *Alox15^-/-^* and *Alox12^-/-^*mice form smaller venous thrombi *in vivo* and have a hemostatic defect that can be corrected using eoxPL administration^15^.

White cell generation of eoxPL was also modulated in patients with arterial vascular disease, which showed increased levels of neutrophil 5-HETE-PL, combined with moderate reductions in other isoforms. Decreased 11- and 15-HETE-PE could result from inhibition of white cell COX activity by aspirin, however due to the small numbers a clear effect was not obvious, and furthermore, 15-HETE-PL in leukocytes can also come from eosinophil 15-LOX which is not sensitive to aspirin^14^. Aspirin use was not conclusively seen to alter leukocyte eoxPL, however the use of P2Y12 inhibitors was associated with significant reductions in 12-HETE-PL (Figure 2 H). This may be due to lower levels of contaminating platelets or platelet-derived extracellular vesicles present as platelet:leukocyte aggregates, since it’s known that P2Y12 inhibitors impair their formation^58, 59^. Overall, apart from 12-HETE-PL, the small suppression of HETE-PLs in white cells appears more likely related to disease, unlike for platelets where 12-LOX acyl-12-HETE-PLs 11- and 15-HETE-PLs from COX-1 are directly impacted by aspirin. An indirect impact of P2Y12 inhibitors on isolated white cells from patients may be a reflection of the strong component of inflammation in arterial vascular disease, where a role for white cells in driving atherosclerosis is well established^60–62^.

In a healthy cohort, complex gender and seasonal differences in platelet eoxPL formation were seen. First, platelets from females mount a stronger response to thrombin in relation to eoxPL formation, a finding that is supported by numerous previous studies showing that platelets from females aggregate more robustly to several agonists in vitro^63–65^. This gender difference was largely abrogated in response to aspirin, suggesting that it may in part rely on secondary activation triggered by TXA_2_. A second gender difference was seen for formation of diacyl 12-HETE-PL, which tended to be elevated in males by aspirin, but not females, while no elevation in plasmalogens in either gender was seen when on aspirin. This mirrored the significant elevations seen in male patients in the clinical cohort further evidencing a gender specific effect of aspirin on generation of a subgroup of eoxPL by human platelets.

Unexpectedly, while most eoxPL were relatively stable across a 12-month sampling period, COX-1 and LOX derived eoxPL steadily increased throughout, peaking in spring. Although only a relatively small increase of 1.5-3 fold, it was consistently observed for ∼15 individual lipid species. The seasonal change for COX-1-derived eoxPL was blunted by aspirin. The basis of this fluctuation is unknown, and we found no reports of seasonal variation in platelet activities in the literature.

Limb or STEMI clots were characterized by a strong platelet eoxPL signature, while white cell or non-enzymatic eoxPL dominated carotid thrombi. Despite a similar pathophysiological process of formation under shear stress, the content of the clot could be influenced by other anatomical/pathological factors. For instance, carotid clot content may be contaminated by significant amount of plaque lipid relative to clot. This is due to the nature of the surgical extraction technique which may also explain the lower total amounts of oxPL. Consistent with this, the predominance of 15-HETE-PL in carotid samples which reflect a component of macrophage/foam cell content present in plaque lesions^66^. Free 15-HETE has previously been reported as a component of human carotid plaques, but deemed to be non-enzymatically generated^67^. Our chiral analysis agrees with this, since it indicated relatively similar proportions of S and R stereoisomers in carotid plaque. This is consistent with studies examining the enantiomeric composition of hydroxyoctodecadienoic acids (HODEs) within advanced atherosclerotic plaques^68^. The physiological age of the analysed clots may also contribute differences. STEMI clots were all retrieved within a short time from formation (<3 hours) compared to carotid clots (days/weeks). Thus, the 12S-HETE-PL dominant profile in STEMI clots may relate to an acute burst of platelet activation in the first few hours of thrombosis, compared to the carotid clots where time has passed since formation which allows for clot metabolism to take place by the generation of an organized fibrin scaffold, modification to the cellular components and possible PL degradation^69–71^. This could also contribute to higher observed amounts of HETE-PL seen owing due to a level of inflammatory cell activation in the acute phase^72^. Clots collected from limb embolectomies varied in retrieval time (‘age’) and this may explain the mixed picture observed in the multivariate analysis where despite an overall similar profile to STEMI, at least two of the ‘oldest’ clots were behaving in a way closer to carotid clots as seen on the heatmap. Other factors that may explain the variability may include inter-individual variability, co-morbidity profile, medications, or lifestyle choices. However, the current sample size is not sufficiently powered to investigate these factors in depth.

Last, *Alox15* was seen to be required for murine arterial thrombosis in vivo. These mice are unable to generate the full range of eoxPL in their blood during ex vivo clotting^15^. This suggests that lipids from 12/15-LOX contribute strongly to acute thrombosis, further supporting our previous studies on venous thrombosis and tail bleeding in both *Alox15* and *Alox12*-deficient strains^8, 14^. In our previous studies, the hemostatic defect could be prevented by administration of low doses of HETE-PE containing liposomes supporting the idea that eoxPL deficiency in the mice plays a direct role^8, 13^.

In summary, we show that procoagulant eoxPL generation is altered in platelets and leukocytes from patients with arterial thrombosis, potentially contributing to elevated thrombotic tendency. Aspirin is the most widely prescribed drug globally for secondary vascular prevention^73, 74^, with its primary effect being inhibition of the platelet lipid, thromboxane A2 (TXA_2_)^75^. Clinically, aspirin or P2Y12 inhibitors are more generally used as anti-platelet agents in arterial disease, versus anti-coagulants like warfarin which predominate clinically in venous thrombosis^76^. Here, we found that both aspirin and P2Y12 inhibitors had complex and selective impacts on eoxPL generation in both health and disease, thus their influence on the pro-coagulant membrane now needs to be considered.

## Data Availability

All data in the present study are available upon reasonable request to the authors

## Supplementary Figure Legends

**Supplementary Figure 1:**
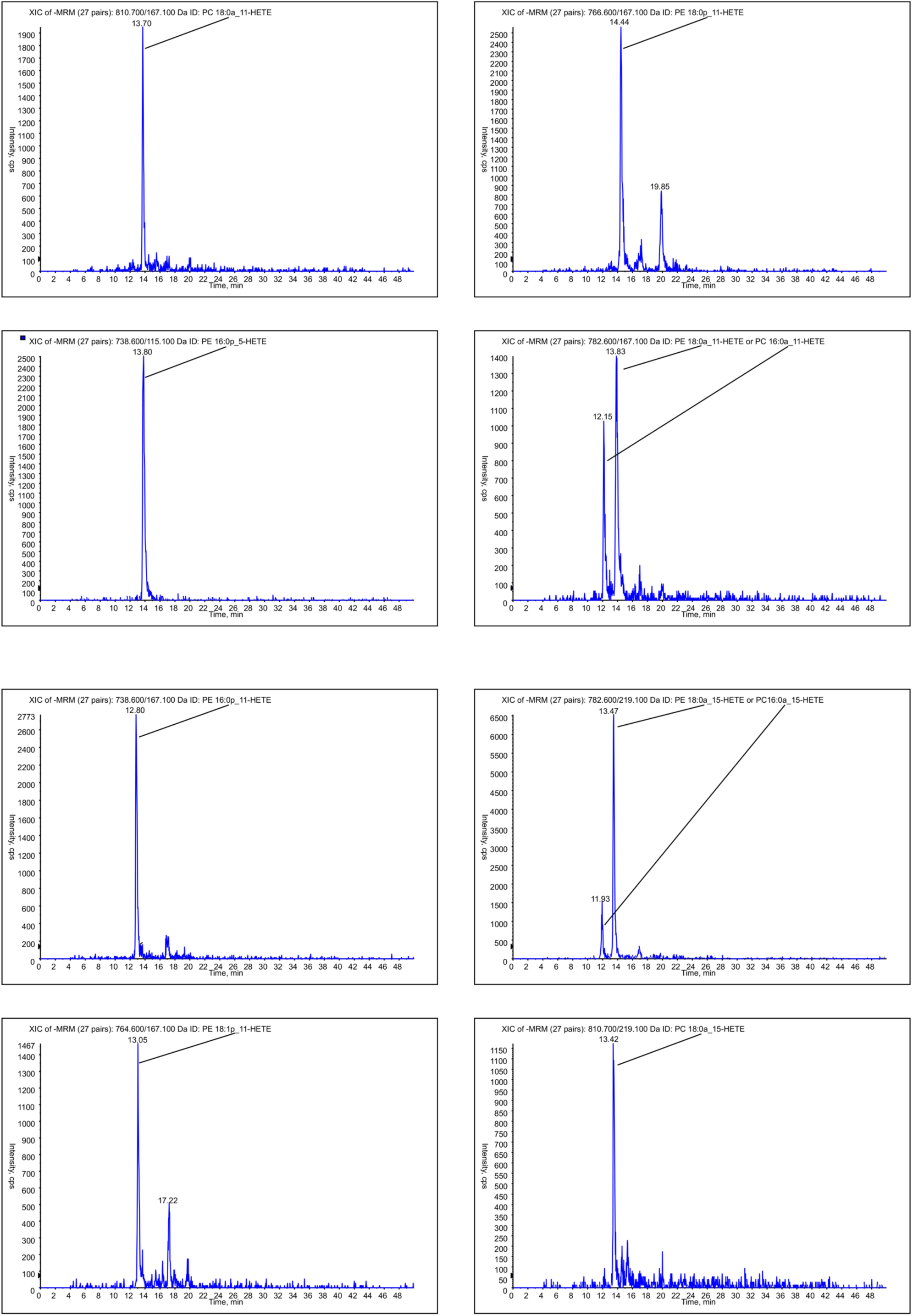
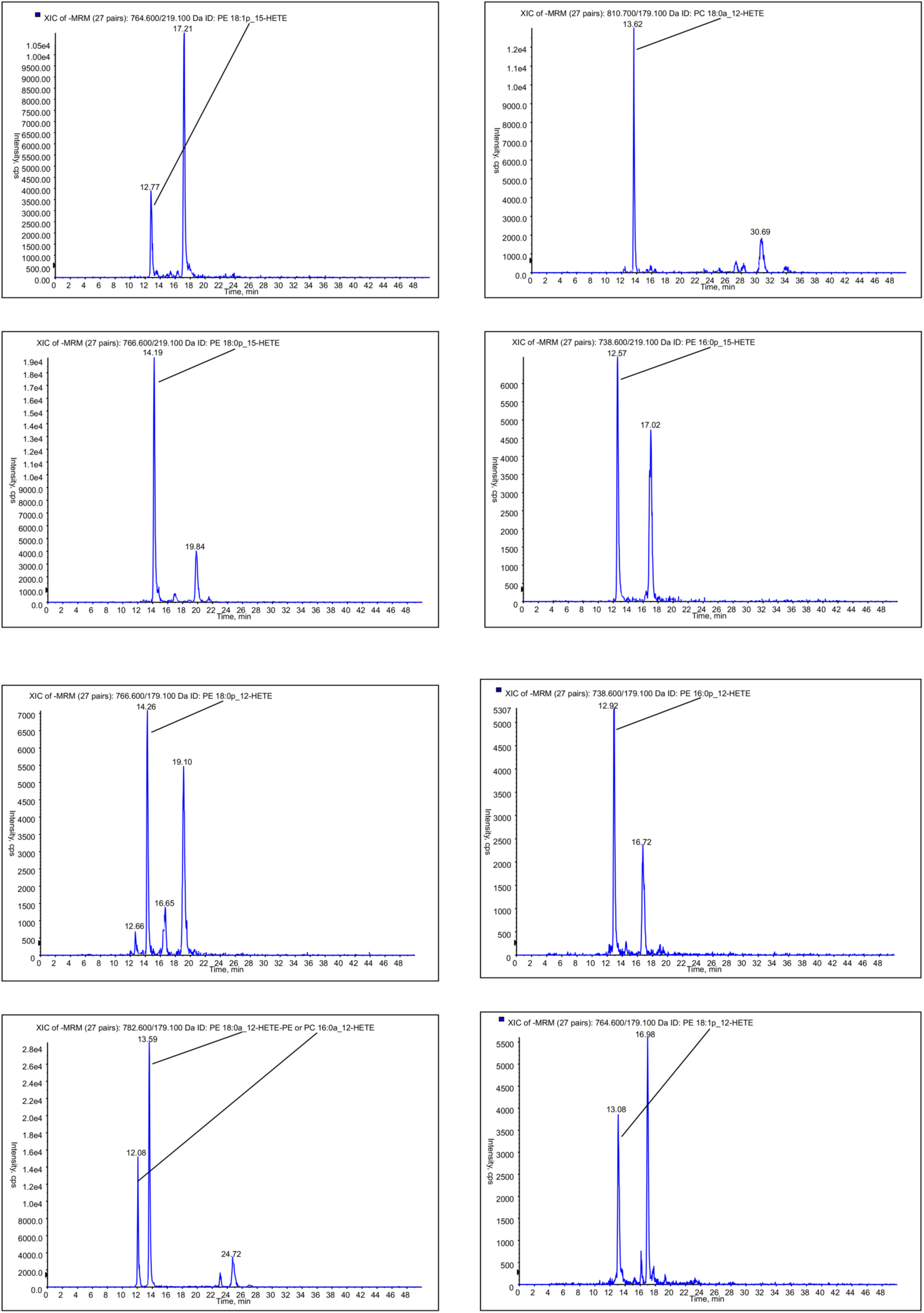
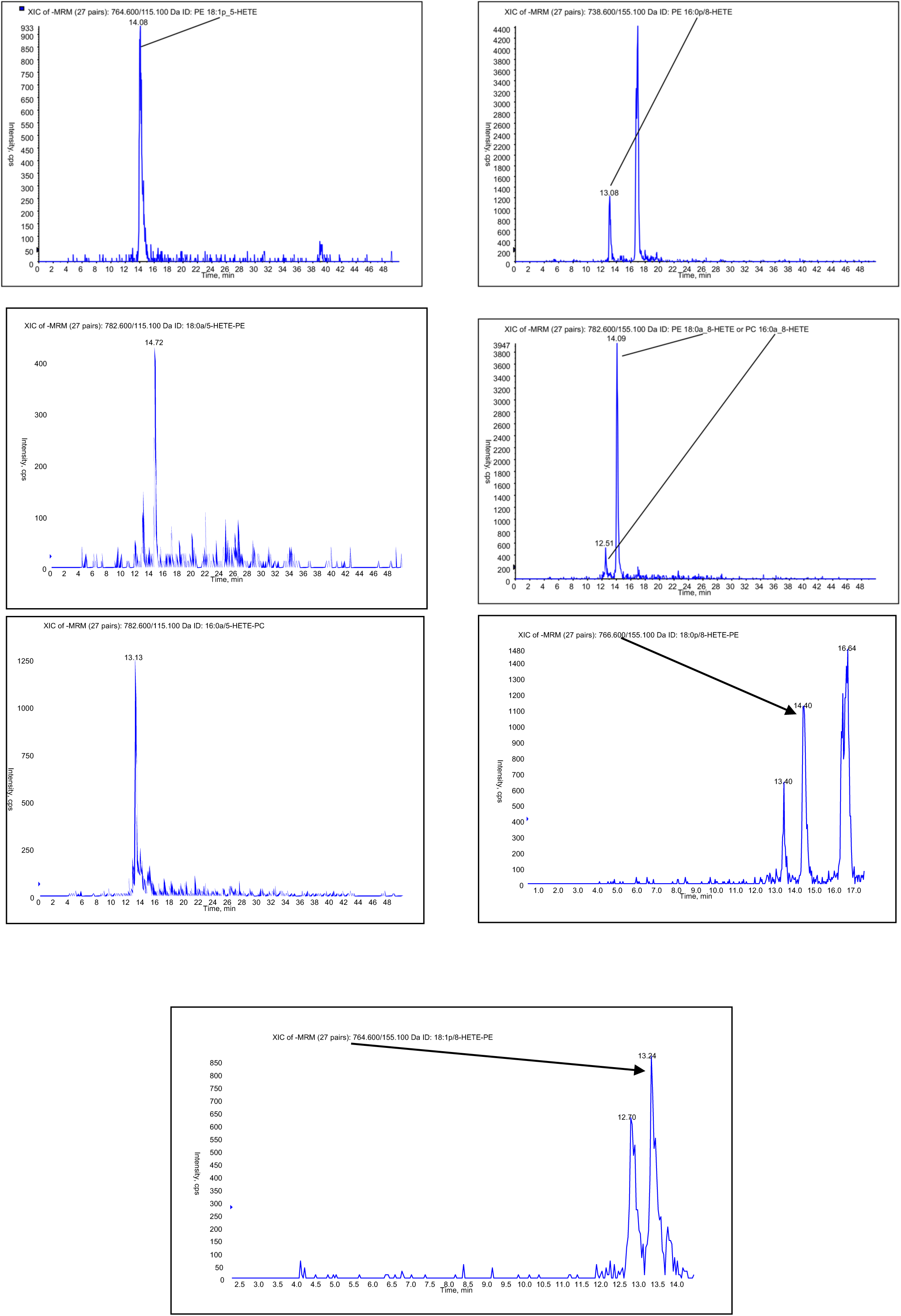
Example chromatograms for all lipids measured in the study. LC/MS/MS was conducted as outlined in Methods.

**Supplementary Figure 2:**
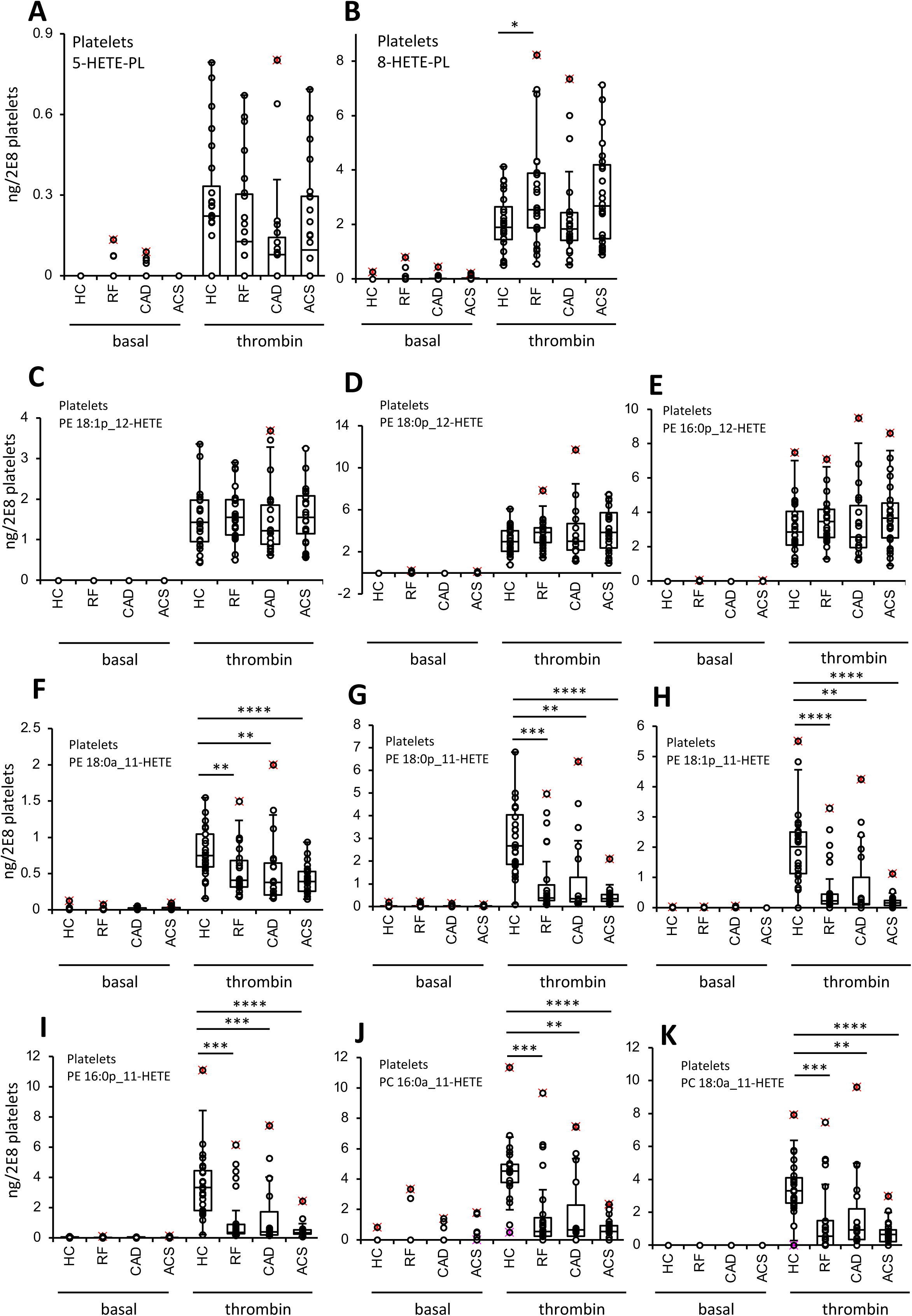
CAD and ACS platelets have reduced COX-1 generated 11-HETE-PL but similar amounts of 12-LOX generated plasmalogen 12-HETE-PL species. Platelets were isolated, activated and lipid extracts then generated and analysed as described in the legend to Figure 1 and Methods. ACS (n=24), CAD (n=19), RF (n=23), HC (n=24). Panels A,B show 5 and 8-HETE-PL respectively. Panels C-E show three plasmalogen 12-HETE-PL lipids. Panels F-K show six individual 11-HETE-PL. Statistical significance was tested with Mann-Whitney-Wilcoxon test for pairwise comparison (* p <0.05, ** p <0.01, *** p <0.001).

**Supplementary Figure 3:**
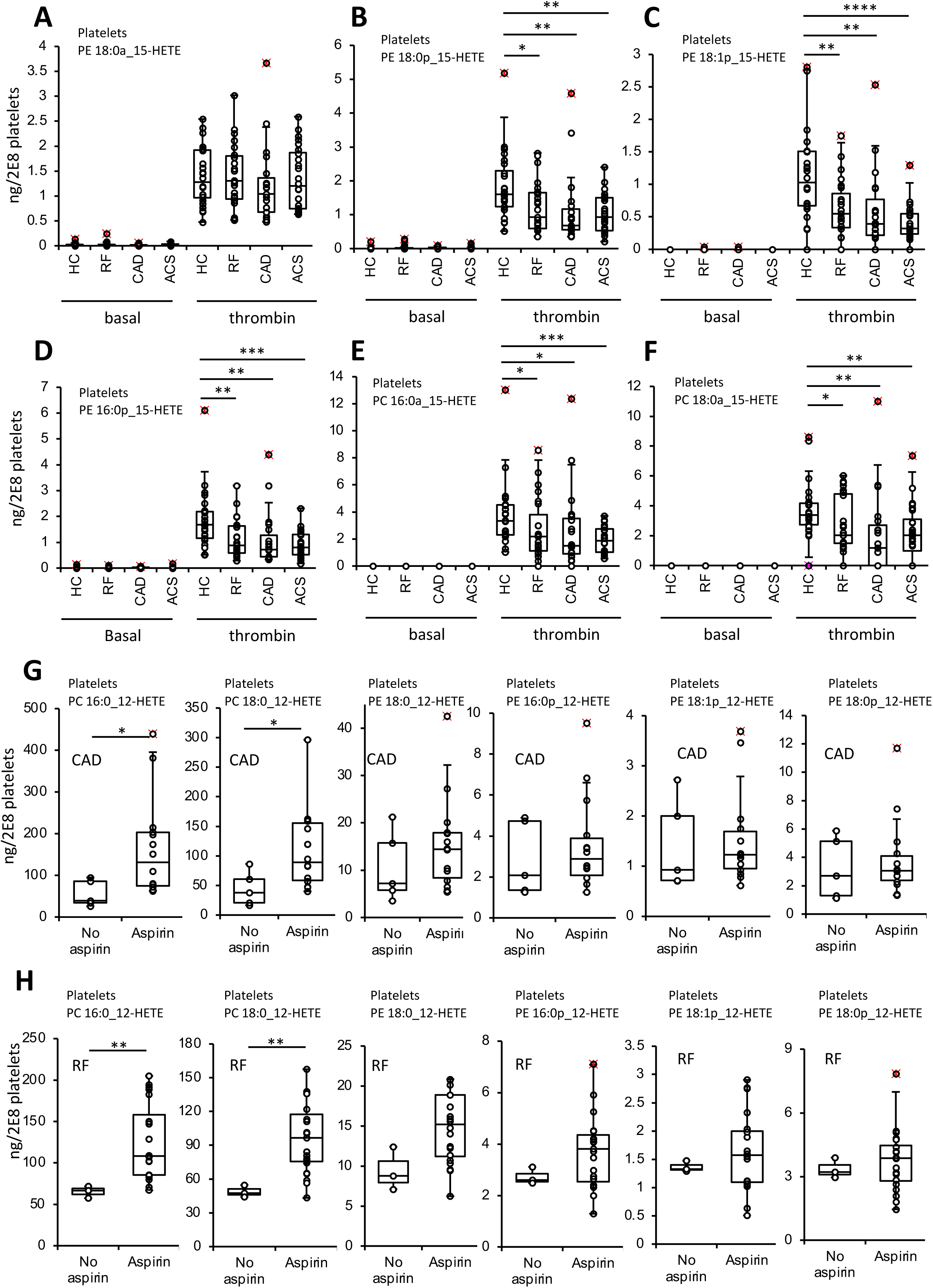
Aspirin reduces COX-1 generated 15-HETE-PL species and increases 12-LOX generated diacyl 12-HETE-PL in thrombin-activated platelets in arterial thrombosis. *Panels A-E: Impact of disease on generation of six plasmalogen 12-HETE-PL lipids*. Platelets were isolated, activated and lipid extracts then generated and analysed as described in the legend to Figure 1 and Methods. ACS (n=24), CAD (n=19), RF (n=23), HC (n=24). *Panels G,H. Aspirin supplementation is associated with elevated levels of diacyl but not plasmalogen 12-HETE-PL in platelets from CAD, and RF patients’ platelets, respectively*. CAD: n = 5, and 14 for no aspirin, and aspirin, respectively, RF: n = 3, and 20 for no aspirin, and aspirin, respectively. Statistical significance was tested with Mann-Whitney-Wilcoxon test for pairwise comparison (* p <0.05, ** p <0.01, *** p <0.001).

**Supplementary Figure 4:**
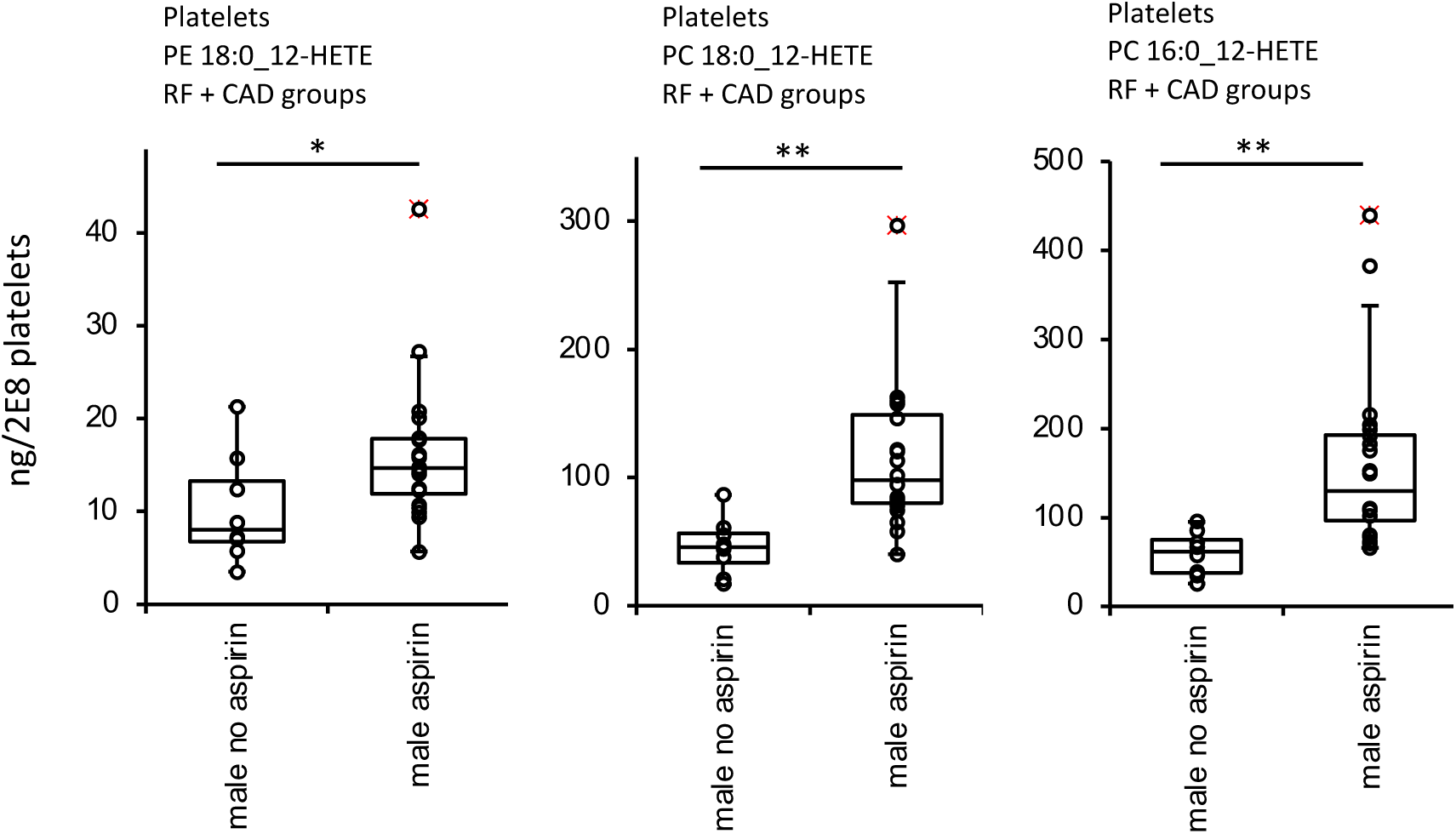
Aspirin increases 12-LOX generated diacyl 12-HETE-PL in thrombin-activated platelets from CAD and RF males. Platelets were isolated, activated and lipid extracts then generated and analysed as described in the legend to Figure 1 and Methods. n = 8, and 20 for no aspirin, and aspirin, respectively. Statistical significance was tested with Mann-Whitney-Wilcoxon test for pairwise comparison (* p <0.05, ** p <0.01, *** p <0.001).

**Supplementary Figure 5:**
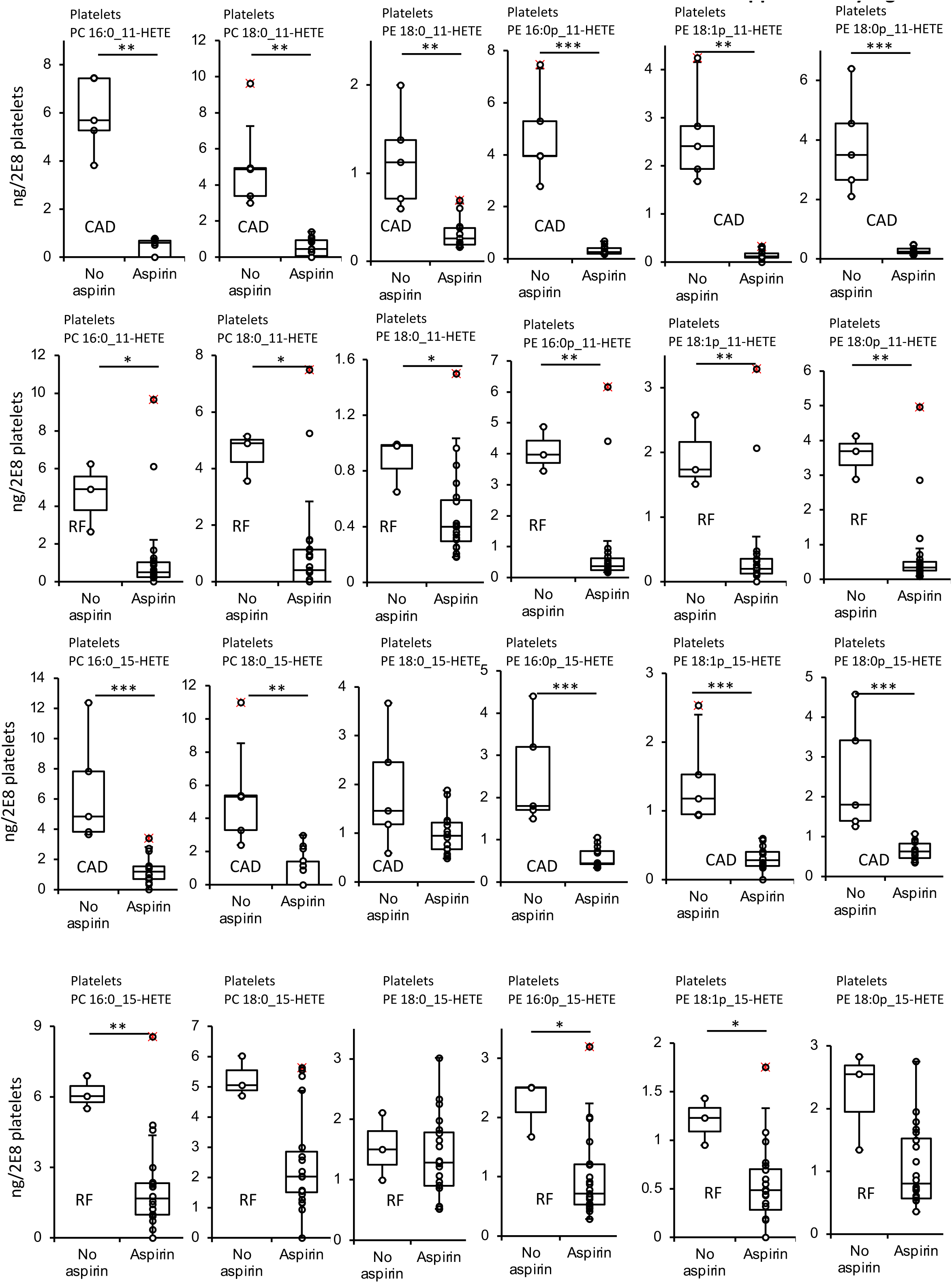
Aspirin reduces COX-1 generated 11- and 15-HETE-PL species in thrombin-activated platelets in arterial thrombosis. Platelets were isolated, activated and lipid extracts then generated and analysed as described in the legend to Figure 1 and Methods. CAD: n = 5, and 14 for no aspirin, and aspirin, respectively, RF: n = 3, and 20 for no aspirin, and aspirin, respectively. *Panel A. CAD, 11-HETE-PLs, Panel B, RF,11-HETE-PL, Panel C, RF, 15-HETE-PE, Panel D, RF, 15-HETE-PE*. Statistical significance was tested with Mann-Whitney-Wilcoxon test for pairwise comparison (* p <0.05, ** p <0.01, *** p <0.001).

**Supplementary Figure 6:**
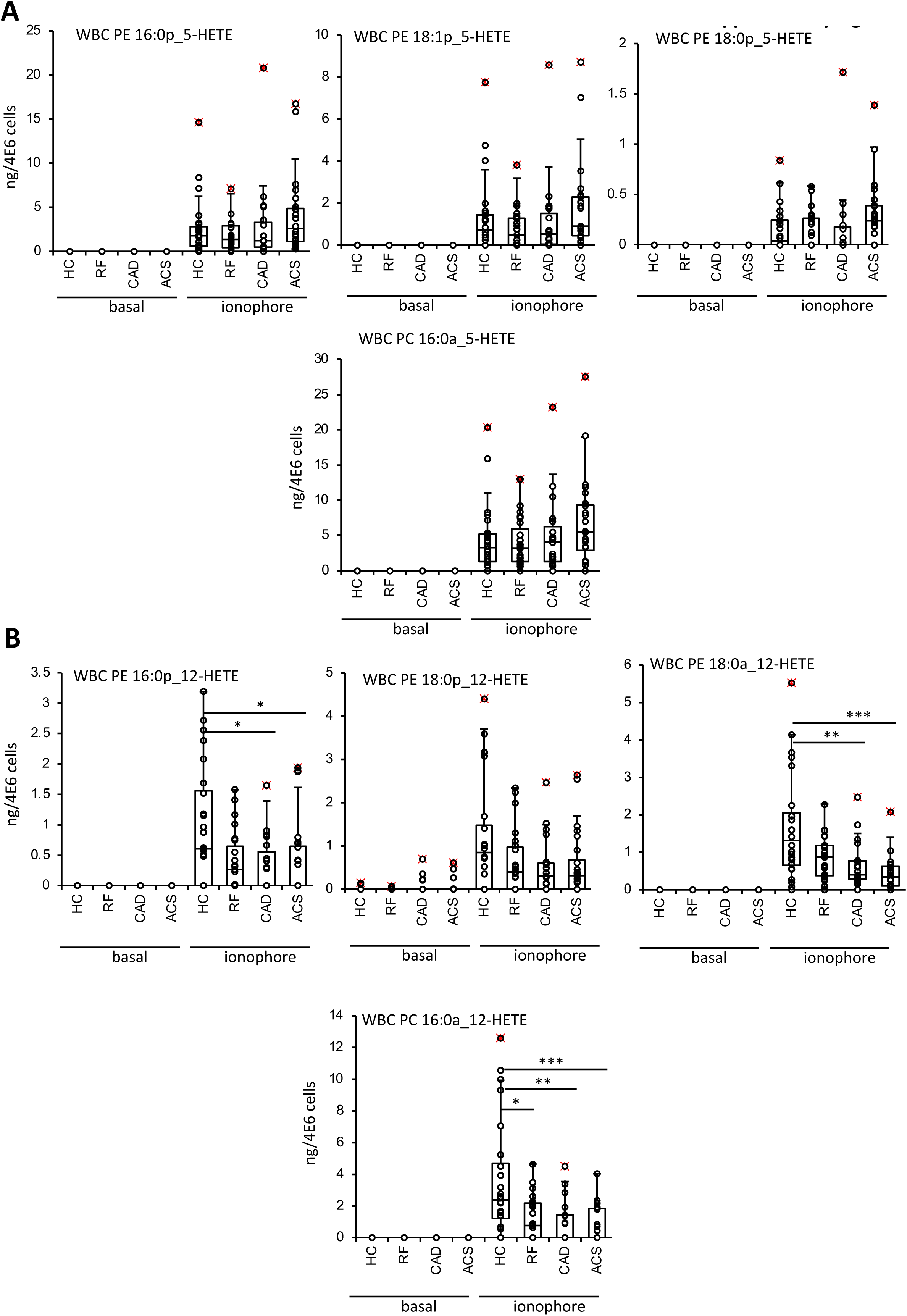
12-HETE-PL generation is reduced in activated leukocytes from patients with arterial thrombosis while 5-HETE-PL are unaffected. Leukocytes were isolated, activated and lipid extracts then generated and analysed as described in the legend to Figure 2 and Methods. *Panel A. 5-HETE levels were relatively unaffected by disease. Panel B. 12-HETE-PLs generated by activated leukocytes are reduced in disease*. ACS (n=24), CAD (n=19), RF (n=23), HC (n=24). Statistical significance was tested with Mann-Whitney-Wilcoxon test for pairwise comparison (*: p <0.05, **: p <0.01, ***:p <0.001, ns or no stars: not significant).

**Supplementary Figure 7:**
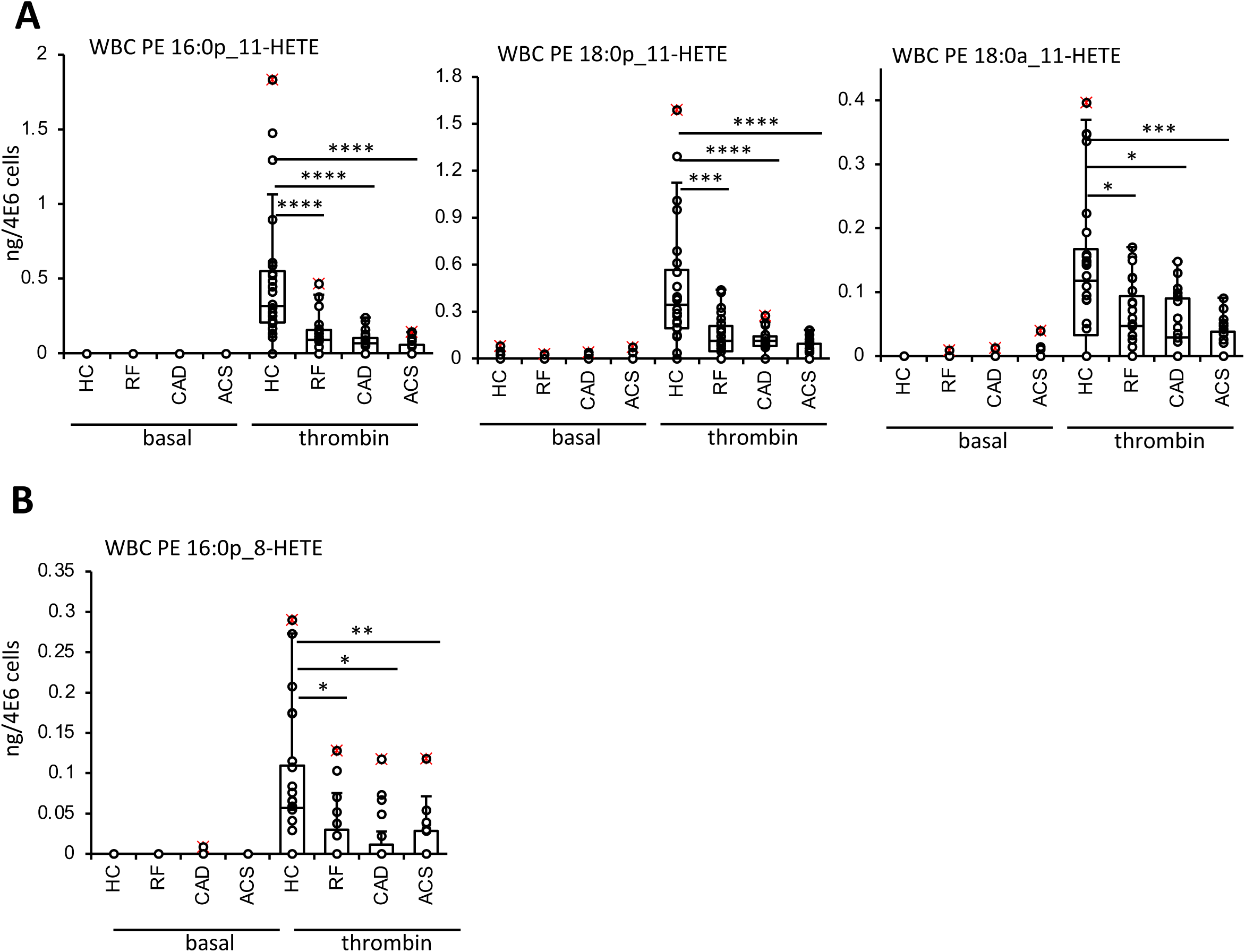
Ionophore-activated leukocytes generate lower levels of 11- and 8-HETE-PL in patients with arterial thrombosis. Leukocytes were isolated, activated and lipid extracts then generated and analysed as described in the legend to Figure 2 and Methods. *Panel A. 11-HETE-PL levels were reduced in platelets from patients with arterial disease. Panel B. 8-HETE-PLs generated by activated leukocytes are reduced in disease* ACS (n=24), CAD (n=19), RF (n=23), HC (n=24). Statistical significance was tested with Mann-Whitney-Wilcoxon test for pairwise comparison (*: p <0.05, **: p <0.01, ***:p <0.001, ns or no stars: not significant).

**Supplementary Figure 8:**
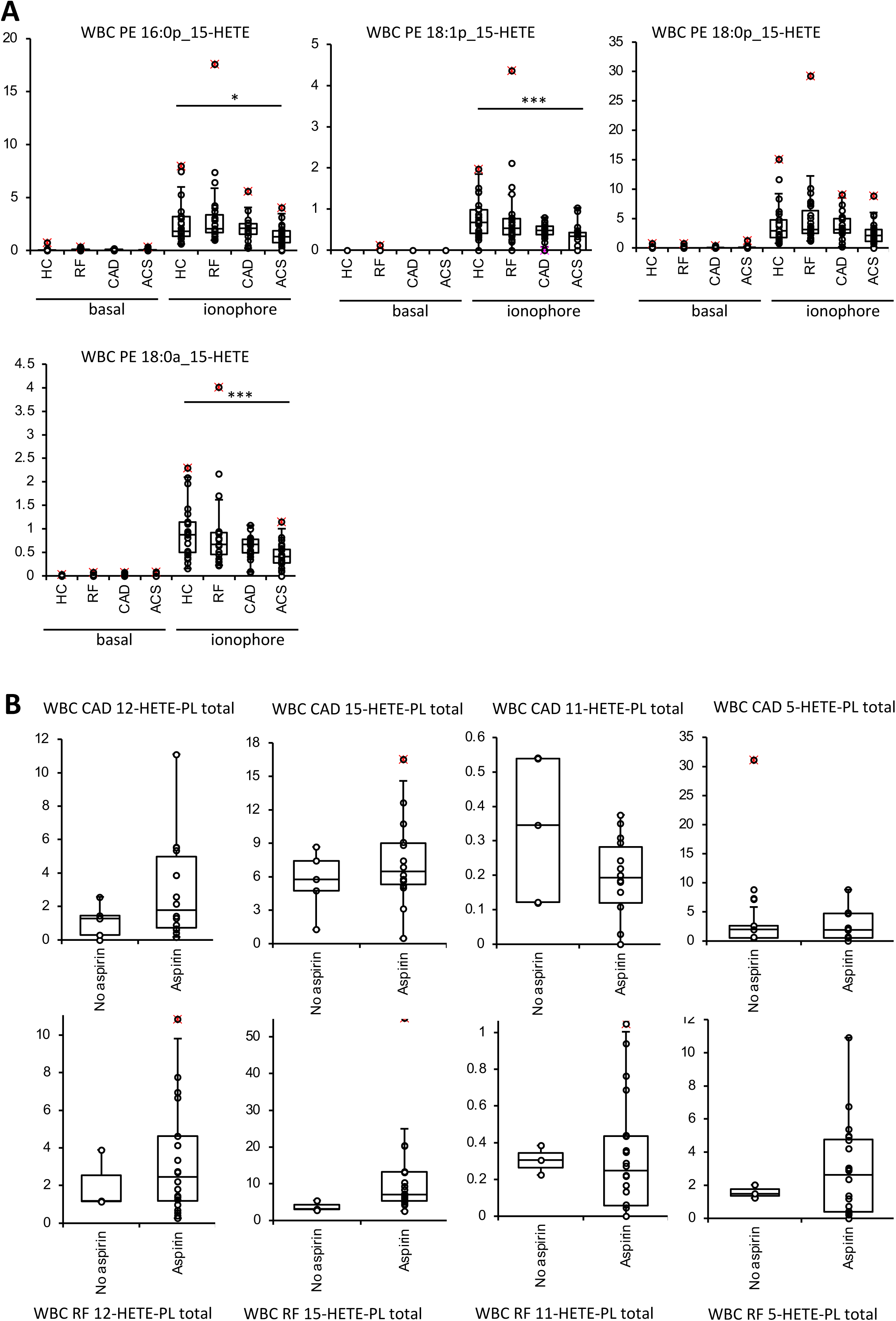
Activated leukocytes from patients with arterial disease generate less 15-HETE-PL, but this is not influenced by aspirin supplementation. Leukocytes were isolated, activated and lipid extracts then generated and analysed as described in the legend to Figure 2 and Methods. *Panel A. 15-HETE-PL levels were reduced in platelets from patients with arterial disease.* ACS (n=24), CAD (n=19), RF (n=23), HC (n=24). Panel B. Aspirin doesn’t influence HETE-PL generation by leukocytes from CAD (upper panels) or RF patients (lower panels). CAD: n = 5, and 14 for no aspirin, and aspirin, respectively, RF: n = 3, and 20 for no aspirin, and aspirin, respectively. Statistical significance was tested with Mann-Whitney-Wilcoxon test for pairwise comparison (* p <0.05, ** p <0.01, *** p <0.001).

**Supplementary Figure 9:**
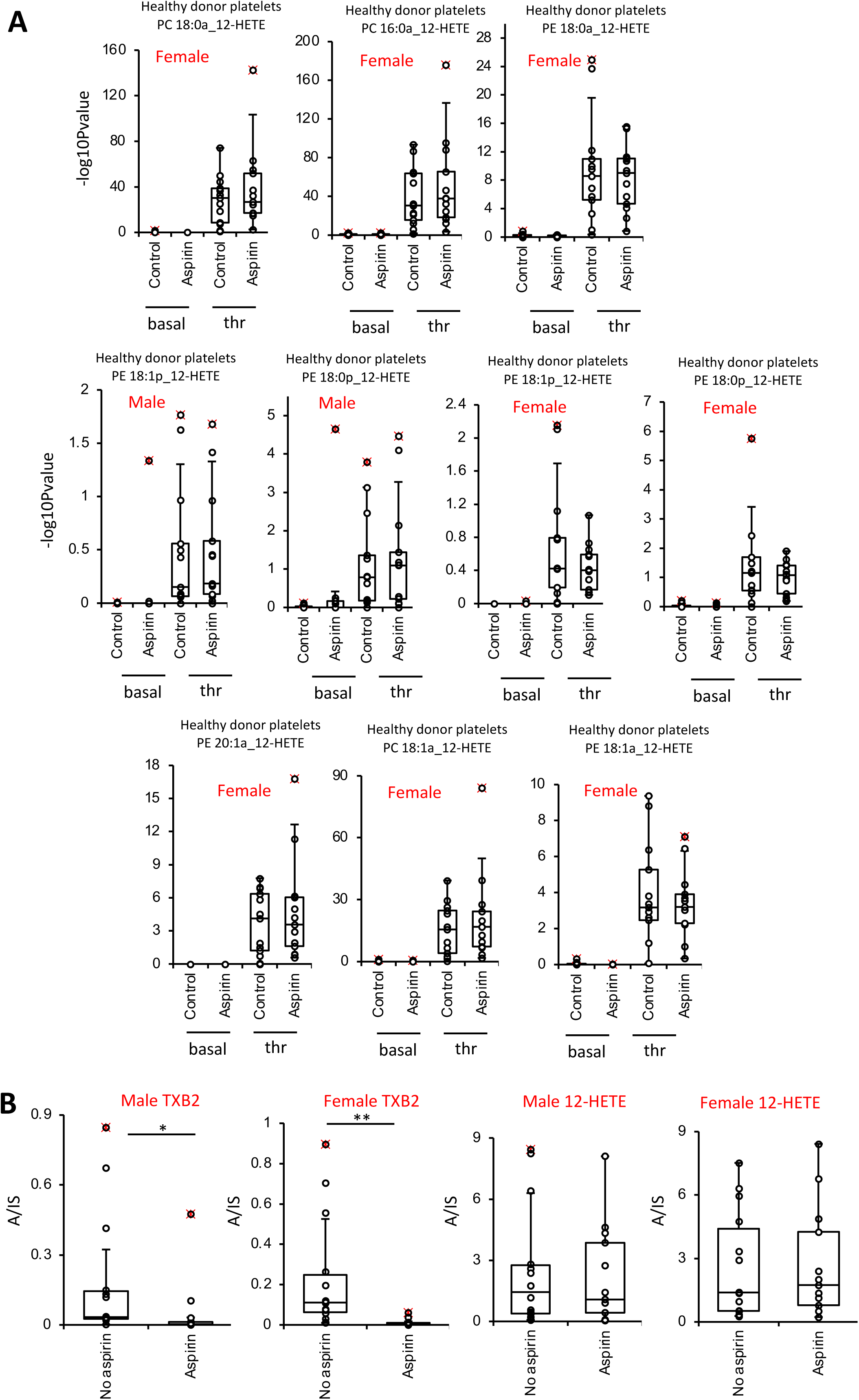
Aspirin supplementation does not influence generation of plasmalogen 12-HETE-PL (males and females), diacyl 12-HETE-PL (females only) or free 12-HETE (males and females). Platelets were isolated from 28 healthy volunteers (n = 14 male, 14 female), activated, and lipids extracted and analysed using LC/MS/MS, as outlined in the legend for Figure 3 and Methods. *Panel A. Box plots showing 12-HETE-PL molecular species in platelets which were not elevated by aspirin supplementation* . Lipids were extracted and analysed using LC/MS/MS as outlined in Methods. *Panel B. Box plots showing that aspirin prevents TXB_2_ generation in males and females while having no impact on free 12-HETE*. Statistical significance was tested with paired T-test (*: p <0.05, **: p <0.01, ***:p <0.001, ns or no stars: not significant).

**Supplementary Table 1.**
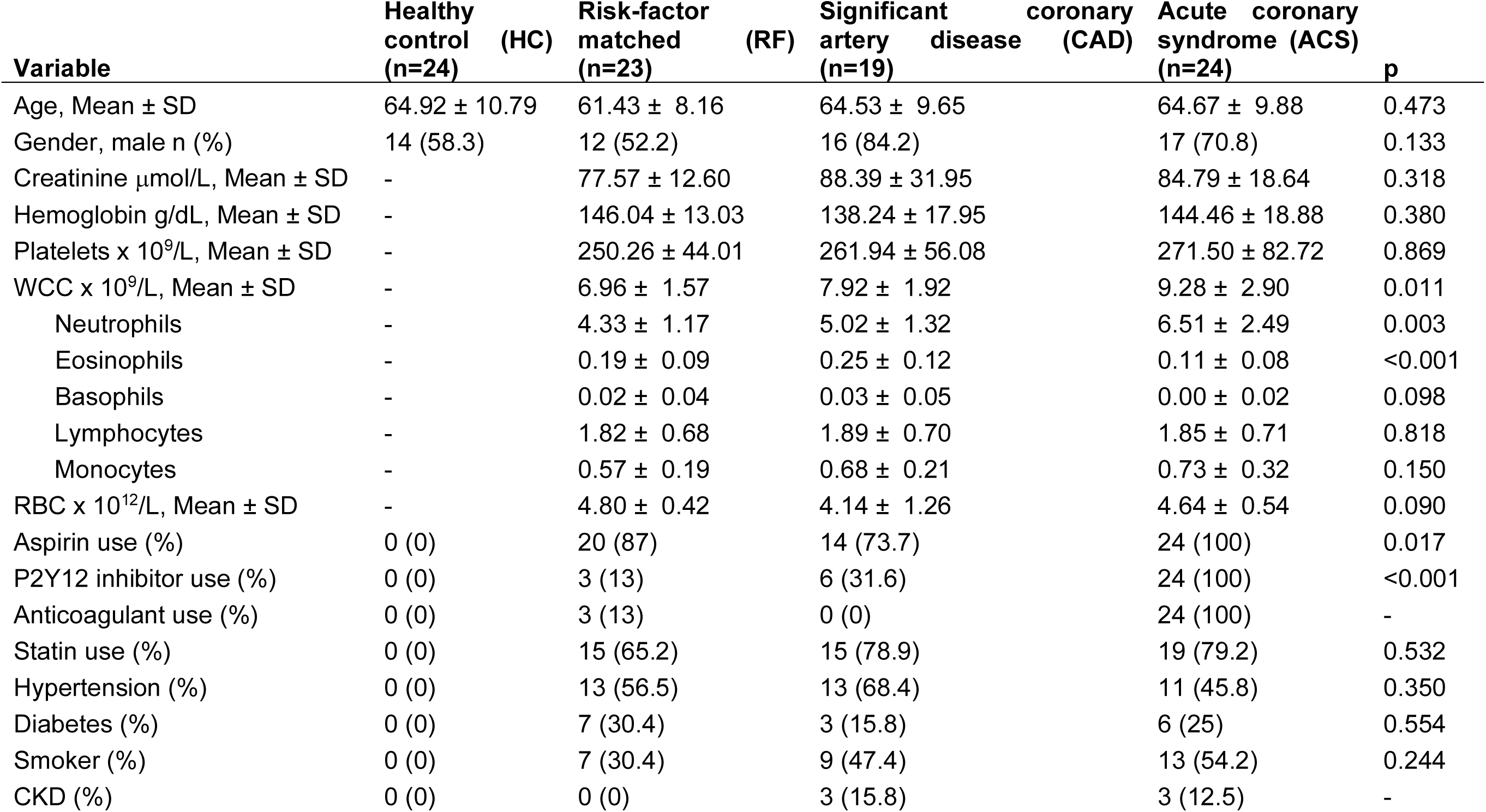
Baseline clinical characteristics of patients recruited in the clinical cohort. (WCC: white cell count, RBC: red blood cell count, P2Y12 inhibitors: clopidogrel, prasugrel or ticagrelor, CKD: chronic kidney disease, SD: standard deviation, p-value tests: Fisher exact (categorical) or Kruskal-Wallis (continuous), p-value comparators: all clinical groups (age, gender) or all except HC for other variables)

**Supplementary Table 2.** Baseline clinical characteristics of extracted clots from patients with atherothrombosis undergoing surgical/interventional clot retrieval. (P2Y12 inhibitors: clopidogrel, prasugrel or ticagrelor, CKD: chronic kidney disease)

| Clot ID | Tissue origin | Age | Gender | Aspirin | P2Y12 inhibitor | Anticoagulant | Statin | Hypertension | Diabetes | Smoker | CKD |
| --- | --- | --- | --- | --- | --- | --- | --- | --- | --- | --- | --- |
| STEMI1 | Left anterior descending artery | 66-70 | Male | Yes | Yes | Warfarin | No | Yes | No | Never | No |
| STEMI2 | Left anterior descending artery | 56-60 | Male | Yes | Yes | Heparin | No | No | No | Never | No |
| STEMI3 | Right coronary artery | 56-60 | Male | Yes | Yes | Heparin | Yes | Yes | No | Ex | No |
| STEMI4 | Left anterior descending artery | 81-85 | Male | Yes | Yes | Heparin | Yes | Yes | No | Never | Yes |
| STEMI5 | Left anterior descending artery | 56-60 | Male | Yes | Yes | Heparin | Yes | Yes | Yes | Never | No |
| STEMI6 | Right coronary artery | 76-80 | Female | Yes | Yes | Heparin | No | No | No | Ex | No |
| Carotid1 | Left Carotid artery | 71-75 | Female | No | No | No | No | No | No | Ex | No |
| Carotid2 | Left Carotid artery | 66-70 | Male | Yes | Yes | No | Yes | Yes | Yes | Ex | No |
| Carotid3 | Right Carotid artery | 71-75 | Female | No | Yes | No | Yes | No | No | Never | No |
| Carotid4 | Left Carotid artery | 61-65 | Male | Yes | Yes | Warfarin | Yes | Yes | No | Never | No |
| Carotid5 | Right Carotid artery | 76-80 | Male | No | Yes | No | Yes | Yes | No | Never | No |
| Carotid6 | Left Carotid artery | 66-70 | Male | No | Yes | No | Yes | Yes | No | Ex | No |
| Carotid7 | Right Carotid artery | 61-65 | Male | Yes | Yes | No | Yes | Yes | No | Never | No |
| Carotid8 | Left Carotid artery | 56-60 | Female | Yes | Yes | No | Yes | No | No | Current | No |
| Limb1 | Right Femoral Embolectomy | 56-60 | Male | Yes | Yes | Heparin | No | No | No | Never | No |
| Limb2 | Left Popliteal (Pop-Ped bypass) | 46-50 | Male | No | Yes | No | Yes | No | No | Current | No |
| Limb3 | Left Femoral (Fem-Pop bypass) | 61-65 | Female | Yes | No | No | No | Yes | Yes | Ex | No |
| Limb4 | Left Popliteal Below-Knee amputation | 56-60 | Male | No | No | Rivaroxaban | No | Yes | Yes | Ex | No |
| Limb5 | Right Brachial Embolectomy | 76-80 | Female | Yes | No | Heparin | No | Yes | No | Never | No |
| Limb6 | Right Popliteal (Fem-Pop bypass with embolectomy) | 61-65 | Male | No | Yes | No | Yes | No | No | Ex | No |

**Supplementary Table 3.** Baseline biochemical and hematological characteristics of extracted clots from patients with atherothrombosis undergoing surgical/interventional clot retrieval. Clot age is the estimated time from diagnosis (clot formation/plaque disruption) to retrieval time. (Creat: serum creatinine μmol/L, Hb: hemoglobin g/dL, Plt: platelet count x 10^9^/L, WCC: total white cell count x 10^9^/L, Neutrophils/Eosinophils/Basophils/Lymphocytes/Monocytes: respective count x 10^9^/L, RBC: red blood cell count x 10^12^/L)

| Clot ID | Clot age | Creat | Hb | Plt | WCC | Neutrophils | Eosinophils | Basophils | Lymphocytes | Mono- | RBC |
| --- | --- | --- | --- | --- | --- | --- | --- | --- | --- | --- | --- |
| STEMI1 | <3hr | 137 | 172 | 168 | 12.1 | 10.7 | 0 | 0 | 0.9 | 0.5 | 5.3 |
| STEMI2 | <3hr | 73 | 128 | 287 | 16.2 | 14.7 | 0.2 | 0 | 0.6 | 0.6 | 4.3 |
| STEMI3 | <3hr | 96 | 149 | 301 | 7.1 | 5.4 | 0.1 | 0 | 1.1 | 0.4 | 4.5 |
| STEMI4 | <3hr | 183 | 137 | 219 | 17 | 14.7 | 0.1 | 0 | 0.9 | 1.2 | 4.8 |
| STEMI5 | <3hr | 92 | 132 | 283 | 14 | 12.5 | 0 | 0 | 0.5 | 1.5 | 4.4 |
| STEMI6 | <3hr | 83 | 120 | 202 | 8.1 | 6.1 | 0 | 0 | 1.4 | 0.5 | 3.9 |
| Carotid1 | 14 days | 65 | 128 | 208 | 5.8 | 3.8 | 0.1 | 0.1 | 1.3 | 0.6 | 3.4 |
| Carotid2 | 20 days | 77 | 130 | 233 | 10.7 | 7.2 | 0.3 | 0.1 | 2.3 | 0.9 | 4.4 |
| Carotid3 | 13 days | 69 | 131 | 425 | 13.1 | 9.5 | 0 | 0 | 2.5 | 1.1 | 3.9 |
| Carotid4 | 9 days | 70 | 133 | 250 | 7.4 | 5.3 | 0.1 | 0 | 1.5 | 0.5 | 4.5 |
| Carotid5 | 6 days | 122 | 154 | 269 | 8.4 | 5.5 | 0.1 | 0 | 1.9 | 0.9 | 5.3 |
| Carotid6 | 25 days | 67 | 129 | 225 | 6 | 4.6 | 0.1 | 0 | 0.9 | 0.4 | 4.8 |
| Carotid7 | 106 days | 82 | 165 | 302 | 5.3 | 3.4 | 0.1 | 0 | 1.2 | 0.6 | 5.1 |
| Carotid8 | 9 days | 80 | 150 | 294 | 8.5 | 5.2 | 0.1 | 0 | 2.7 | 0.6 | 4.9 |
| Limb1 | 3 days | 69 | 125 | 501 | 10.5 | 7.5 | 0.1 | 0.1 | 2.2 | 0.8 | 4.0 |
| Limb2 | 4 days | 95 | 174 | 306 | 7.1 | 5.3 | 0.1 | 0 | 1.2 | 0.4 | 6.0 |
| Limb3 | >6 weeks | 59 | 99 | 459 | 13.6 | 10.7 | 0 | 0 | 1.6 | 1.2 | 3.0 |
| Limb4 | 6 days | 70 | 83 | 163 | 8.3 | 6.1 | 0 | 0 | 1.5 | 0.6 | 2.7 |
| Limb5 | 16 days | 91 | 162 | 309 | 7.8 | 4.1 | 0.1 | 0.1 | 3.0 | 0.6 | 5.3 |
| Limb6 | 15 days | 63 | 179 | 313 | 9.2 | 6.3 | 0.5 | 0.1 | 1.0 | 1.3 | 5.8 |

**Supplementary Table 4.** MRM transitions for eoxPL analysed from the healthy cohort.

| Name (ID) | Precursor m/z (Q1) | Product ion m/z (Q3) | RT |
| --- | --- | --- | --- |
| PE 16:0p_HETE | 738.6 | 319.2 | 12.59 |
| PE 16:0a_HETE | 754.6 | 319.2 | 12.02 |
| PE 16:0p_20:4;2O | 754.6 | 335.2 | 10.95 |
| PE 16:0p_HDoHE | 762.6 | 343.2 | 12.02 |
| PE 18:1p_12-HETE | 764.6 | 179.1 | 12.90 |
| PE 18:0p_20:5;O | 764.6 | 317.2 | 14.22 |
| PE 16:0p_22:5;O | 764.6 | 345.2 | 12.50 |
| PE 18:0p_12-HETE | 766.6 | 179.1 | 14.10 |
| PE 18:0p_15-HETE | 766.6 | 219.1 | 13.63 |
| PE 16:0e_22:5;O | 766.6 | 345.2 | 12.92 |
| PE 16:0p_22:4;O | 766.6 | 347.2 | 13.28 |
| PE 16:0a_20:4;2O | 770.6 | 335.2 | 10.95 |
| PE 16:0p_20:4;3O | 770.6 | 351.2 | 9.40 |
| PE 16:0p_Di-EHEDA | 770.6 | 351.2 | 10.03 |
| PE 18:2a_HETE | 778.6 | 319.2 | 11.18 |
| PE 16:0a_HDOHE | 778.6 | 343.2 | 11.42 |
| PE 18:1a_12-HETE | 780.6 | 179.1 | 12.25 |
| PE 18:0a_20:5;O | 780.6 | 317.2 | 16.08 |
| PE 18:1p_20:4;2O | 780.6 | 335.2 | 10.93 |
| PE 16:0a_22:5;O | 780.6 | 345.2 | 11.98 |
| PE 16:0p_22:5;2O | 780.6 | 361.2 | 10.93 |
| PE 18:0a_12-HETE | 782.6 | 179.1 | 13.41 |
| PC 16:0a_12-HETE | 782.6 | 179.1 | 11.79 |
| PE 18:0p_HDOHE | 790.6 | 343.2 | 13.72 |
| PE 18:1p_22:5;O | 790.6 | 345.2 | 12.91 |
| PE 18:0p_22:5;O | 792.6 | 345.2 | 14.24 |
| PE 18:1p_22:4;O | 792.6 | 347.2 | 13.90 |
| PE 20:0p_HETE | 794.6 | 319.2 | 13.23 |
| PE 18:0p_22:4;O | 794.6 | 347.2 | 15.32 |
| PE 18:1p_20:4;3O | 796.6 | 351.2 | 9.80 |
| PE 18:0p_20:4;3O | 798.6 | 351.2 | 11.14 |
| PE 18:1p_Di-EHEDA | 796.6 | 351.2 | 10.53 |
| PE 18:0p_Di-EHEDA | 798.6 | 351.2 | 11.73 |
| PE 18:1a_HDoHE | 804.7 | 343.2 | 12.06 |
| PC 18:2a_HETE | 806.7 | 319.2 | 11.18 |
| PC 16:0a_HDOHE | 806.7 | 343.2 | 11.61 |
| PE 18:0a_HDOHE | 806.7 | 343.2 | 13.26 |
| PE 18:1a_22:5;O | 806.7 | 345.2 | 12.29 |
| PE 20:1a_12-HETE | 808.7 | 179.1 | 12.16 |
| PC 18:1a_12-HETE | 808.7 | 179.2 | 12.17 |
| PC 16:0a_22:5;O | 808.7 | 345.2 | 11.89 |
| PE 18:0a_22:5;O | 808.7 | 345.2 | 13.51 |
| PE 18:1a_22:4;O | 808.7 | 347.2 | 13.23 |
| PC 18:0a_12-HETE | 810.7 | 179.1 | 13.41 |
| PE 18:0a_20:4;3O | 814.7 | 351.2 | 10.35 |
| PE 18:0a_Di-EHEDA | 814.7 | 351.2 | 11.00 |
| PE 18:0p_20:4;2O | 782.6 | 335.2 | 13.28 |
| PE 16:0a_22:4;O | 782.6 | 347.2 | 12.80 |
| DMPC (PC 14:0/14:0) | 662.5 | 227.1 | 13.63 |
| DMPE (PE 14:0/14:0) | 634.5 | 227.1 | 13.98 |

**Supplementary Table 5.**
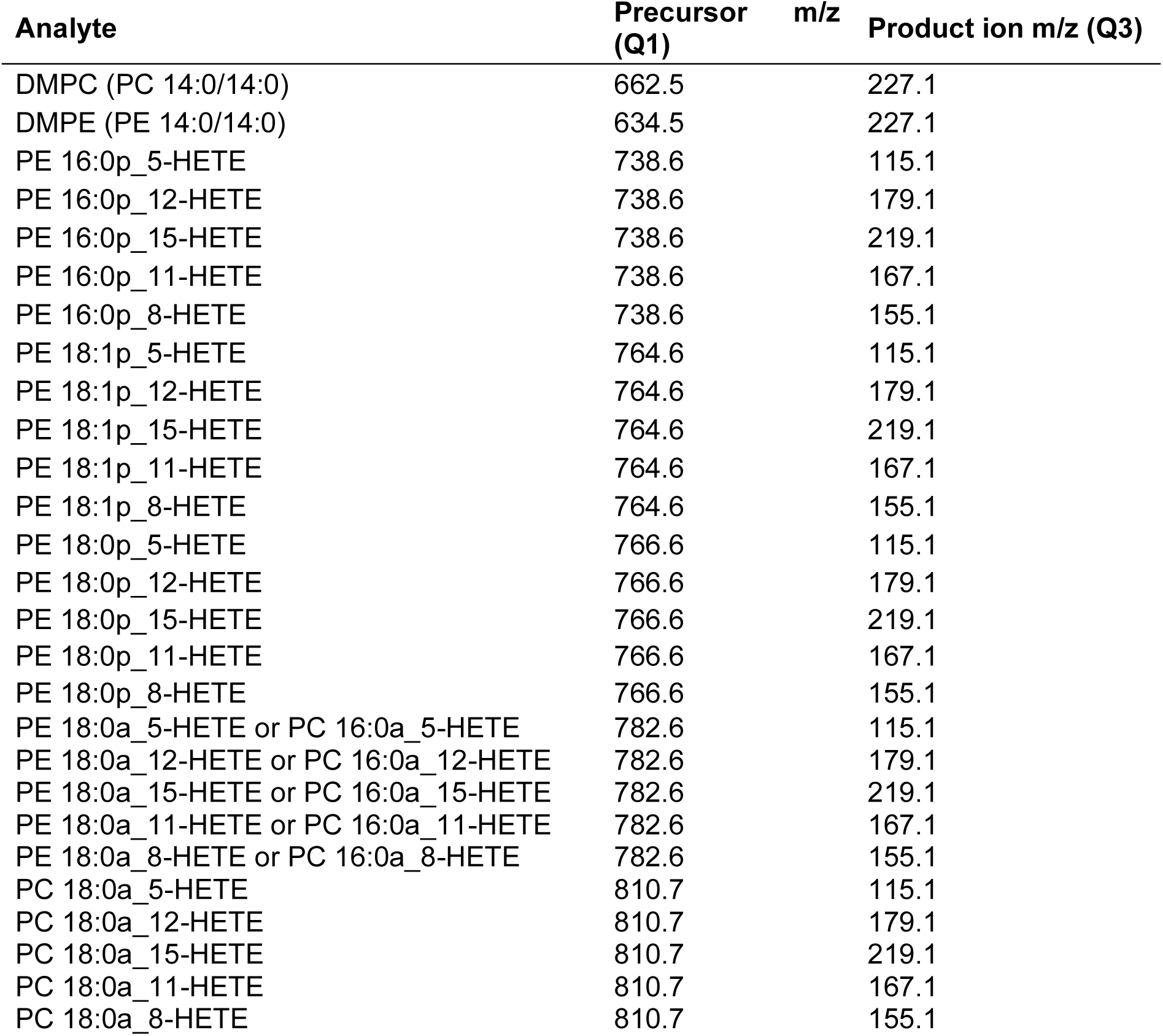
MRM transitions for the oxPL analysed from the clinical cohort and clot samples.

**Supplementary Table 6.** MRM transitions for eicosanoids used for LC/MS/MS.

| Analyte | MRM transition | DP (V) | CE (V) | CXP (V) |
| --- | --- | --- | --- | --- |
| 12-HETE | 319.2→179.1 | -65 | -18 | -12 |
| TxB2 | 369.1→169.1 | -60 | -22 | -12 |
| 12-HETE-d8 | 327.2→184.1 | -60 | -20 | -12 |
| TxB2-d4 | 373.3→173.3 | -55 | -22 | -10 |

**Supplementary Table 7.** Multiple reaction monitoring (MRM) transitions for chiral analysis of hydrolyzed HETE stereoisomers.

| Analyte | MRM transition | DP (V) | CE (V) | CXP (V) |
| --- | --- | --- | --- | --- |
| 15-HETE | 319.2→219.1 | -70 | -20 | -13 |
| 12-HETE | 319.2→179.1 | -75 | -22 | -9 |
| 11-HETE | 319.2→167.1 | -75 | -24 | -1 |
| 8-HETE | 319.2→155.1 | -70 | -22 | -9 |
| 5-HETE | 319.2→115.1 | -70 | -22 | -7 |
| 12(S)-HETE-d8 | 327.2→184.1 | -80 | -22 | -11 |

**Supplementary Table 8.** Demographics for individual patients for aspirin and P2Y12 inhibitors.

| Patient Group | Age | Gender | Aspirin | P2Y12 |  | Patient Group | Age | Gender | Aspirin | P2Y12 |
| --- | --- | --- | --- | --- | --- | --- | --- | --- | --- | --- |
| ACS | 56-60 | Male | Yes | Yes |  | HC | 51-55 | Male | No | No |
| ACS | 66-70 | Female | Yes | Yes |  | HC | 51-55 | Male | No | No |
| ACS | 71-75 | Male | Yes | Yes |  | HC | 71-75 | Female | No | No |
| ACS | 56-60 | Male | Yes | Yes |  | HC | 81-85 | Male | No | No |
| ACS | 51-55 | Male | Yes | Yes |  | HC | 66-70 | Male | No | No |
| ACS | 51-55 | Male | Yes | Yes |  | HC | 46-50 | Female | No | No |
| ACS | 76-80 | Male | Yes | Yes |  | HC | 71-75 | Male | No | No |
| ACS | 66-70 | Female | Yes | Yes |  | HC | 61-65 | Female | No | No |
| ACS | 51-55 | Male | Yes | Yes |  | HC | 61-65 | Male | No | No |
| ACS | 51-55 | Male | Yes | Yes |  | HC | 51-55 | Male | No | No |
| ACS | 61-65 | Male | Yes | Yes |  | HC | 61-65 | Female | No | No |
| ACS | 71-75 | Male | Yes | Yes |  | HC | 76-80 | Male | No | No |
| ACS | 71-75 | Female | Yes | Yes |  | HC | 76-80 | Female | No | No |
| ACS | 61-65 | Female | Yes | Yes |  | HC | 41-45 | Female | No | No |
| ACS | 51-55 | Male | Yes | Yes |  | HC | 66-70 | Male | No | No |
| ACS | 46-50 | Male | Yes | Yes |  | HC | 66-70 | Female | No | No |
| ACS | 61-65 | Male | Yes | Yes |  | HC | 76-80 | Male | No | No |
| ACS | 71-75 | Male | Yes | Yes |  | HC | 61-65 | Male | No | No |
| ACS | 56-60 | Male | Yes | Yes |  | HC | 81-85 | Male | No | No |
| ACS | 76-80 | Male | Yes | Yes |  | HC | 66-70 | Female | No | No |
| ACS | 56-60 | Female | Yes | Yes |  | HC | 66-70 | Female | No | No |
| ACS | 76-80 | Female | Yes | Yes |  | HC | 56-60 | Female | No | No |
| ACS | 71-75 | Female | Yes | Yes |  | HC | 61-65 | Male | No | No |
| ACS | 76-80 | Male | Yes | Yes |  | HC | 51-55 | Male | No | No |
| CAD | 71-75 | Male | No | Yes |  | RF | 71-75 | Female | Yes | No |
| CAD | 66-70 | Male | Yes | No |  | RF | 51-55 | Female | Yes | No |
| CAD | 51-55 | Female | Yes | Yes |  | RF | 76-80 | Female | Yes | No |
| CAD | 66-70 | Female | Yes | No |  | RF | 61-65 | Female | Yes | No |
| CAD | 46-50 | Male | Yes | No |  | RF | 61-65 | Male | No | No |
| CAD | 36-40 | Female | Yes | No |  | RF | 46-50 | Female | Yes | No |
| CAD | 71-75 | Male | Yes | No |  | RF | 71-75 | Female | Yes | No |
| CAD | 61-65 | Male | No | Yes |  | RF | 56-60 | Male | Yes | No |
| CAD | 66-70 | Male | No | Yes |  | RF | 56-60 | Male | Yes | Yes |
| CAD | 71-75 | Male | No | No |  | RF | 51-55 | Male | Yes | No |
| CAD | 56-60 | Male | Yes | No |  | RF | 66-70 | Male | Yes | Yes |
| CAD | 71-75 | Male | Yes | No |  | RF | 66-70 | Male | No | No |
| CAD | 56-60 | Male | Yes | No |  | RF | 56-60 | Female | Yes | No |
| CAD | 61-65 | Male | Yes | No |  | RF | 51-55 | Male | Yes | No |
| CAD | 61-65 | Male | Yes | No |  | RF | 66-70 | Male | Yes | No |
| CAD | 71-75 | Male | Yes | No |  | RF | 51-55 | Female | Yes | No |
| CAD | 61-65 | Male | Yes | No |  | RF | 51-55 | Male | Yes | No |
| CAD | 66-70 | Male | No | Yes |  | RF | 66-70 | Male | Yes | Yes |
| CAD | 71-75 | Male | Yes | Yes |  | RF | 66-70 | Male | No | No |
|  |  |  |  |  |  | RF | 56-60 | Female | Yes | No |
|  |  |  |  |  |  | RF | 56-60 | Female | Yes | No |
|  |  |  |  |  |  | RF | 61-65 | Female | Yes | No |
|  |  |  |  |  |  | RF | 46-50 | Male | Yes | No |

**Supplementary Table 9.** Change in eoxPL amounts generated by thrombin-activated platelets (compared with resting platelets) in participants from a healthy cohort.

| **Lipid Name** | **Mean A/IS**<br>**(Resting)** | **Mean A/IS**<br>**(Thrombin)** | **Fold change**<br>**(thrombin/resting)** | **p-value** |
| --- | --- | --- | --- | --- |
| PE 18:0a_12-HETE | 0.0011 | 0.05 | 45.45 | 2.6E-16 |
| PE 20:1a_12-HETE | 0.00011 | 0.021 | 190.91 | 1.9E-11 |
| PE 18:1p_12-HETE | 0.000011 | 0.0041 | 372.73 | 2.3E-11 |
| PC 18:1a_12-HETE | 0.0004 | 0.039 | 97.50 | 4.7E-11 |
| PC 18:0a_12-HETE | 0.00072 | 0.085 | 118.06 | 5.5E-11 |
| PE 18:1a_12-HETE | 0.00019 | 0.019 | 100.00 | 7.7E-11 |
| PE 18:0p_12-HETE | 0.00027 | 0.0097 | 35.93 | 1.4E-10 |
| PC 16:0a_12-HETE | 0.0027 | 0.1 | 37.04 | 2.9E-10 |
| PE 16:0p_HETE | 0.0032 | 0.21 | 65.63 | 2.6E-16 |
| PE 16:0a_HETE | 0.0047 | 0.16 | 34.04 | 2.6E-16 |
| PE 18:2a_HETE | 0.0013 | 0.024 | 18.46 | 2.6E-16 |
| PE 20:0p_HETE | 0.00013 | 0.027 | 207.69 | 6.8E-11 |
| PC 18:2a_HETE | 0.00069 | 0.059 | 85.51 | 1.4E-10 |
| PE 18:0p_DiEHEDA | 0.0018 | 0.026 | 14.44 | 5.2E-16 |
| PE 16:0p_20:4;3O | 0.0012 | 0.018 | 15.00 | 5.2E-16 |
| PE 18:0p_20:4;3O | 0.0041 | 0.037 | 9.02 | 1.8E-15 |
| PE 18:0a_20:4;3O | 0.0026 | 0.016 | 6.15 | 3.1E-15 |
| PE 18:0a_DiEHEDA | 0.0013 | 0.0065 | 5.00 | 1.5E-12 |
| PE 18:0p_15-HETE | 0.001 | 0.0063 | 6.30 | 3.1E-12 |
| PE 18:1p_20:4;3O | 0.00026 | 0.014 | 53.85 | 1.2E-10 |
| PE 18:1p_DiEHEDA | 0.00046 | 0.016 | 34.78 | 1.6E-10 |
| PE 16:0p_DiEHEDA | 0.001 | 0.024 | 24.00 | 1.9E-10 |
| PE 18:0p_22:5;O | 0.0087 | 0.35 | 40.23 | 2.6E-16 |
| PC 16:0a_HDoHE | 0.0024 | 0.045 | 18.75 | 2.6E-16 |
| PE 18:0a_HDoHE | 0.0031 | 0.14 | 45.16 | 2.6E-16 |
| PE 16:0p_HDoHE | 0.0025 | 0.12 | 48.00 | 2.6E-16 |
| PE 18:0a_22:5;O | 0.0048 | 0.22 | 45.83 | 5.2E-16 |
| PE 16:0p_22:5;O | 0.0065 | 0.3 | 46.15 | 5.2E-16 |
| PE 16:0p_22:4;O | 0.0099 | 0.35 | 35.35 | 5.2E-16 |
| PE 16:0e_22:5;O | 0.00021 | 0.027 | 128.57 | 3.6E-11 |
| PE 18:1p_22:5;O | 0.00065 | 0.059 | 90.77 | 4.1E-11 |
| PE 18:1a_22:5;O | 0.00061 | 0.059 | 96.72 | 5.2E-11 |
| PE 18:1a_22:4;O | 0.00063 | 0.054 | 85.71 | 6E-11 |
| PE 18:1p_22:4;O | 0.00063 | 0.053 | 84.13 | 8.5E-11 |
| PE 18:1a_HDoHE | 0.00055 | 0.037 | 67.27 | 1.1E-10 |
| PE 16:0a_22:4;O | 0.0011 | 0.074 | 67.27 | 1.2E-10 |
| PE 18:0p_HDoHE | 0.0018 | 0.13 | 72.22 | 1.3E-10 |
| PE 18:0p_22:4;O | 0.0041 | 0.21 | 51.22 | 1.4E-10 |
| PE 16:0a_22:5;O | 0.0016 | 0.098 | 61.25 | 1.4E-10 |
| PC 16:0a_22:5;O | 0.0012 | 0.062 | 51.67 | 1.6E-10 |
| PE 16:0a_HDoHE | 0.00079 | 0.037 | 46.84 | 1.6E-10 |
| PE 16:0p_22:5;2O | 0.0054 | 0.017 | 3.15 | 7.5E-10 |
| PE 18:1p_20:4;2O | 0.0037 | 0.0087 | 2.35 | 0.000001 |
| PE 16:0a_20:4;2O | 0.0019 | 0.0045 | 2.37 | 0.00021 |
| PE 16:0p_20:4;2O | 0.0087 | 0.012 | 1.38 | 0.027 |
| PE 18:0p_20:5;O | 0.012 | 0.017 | 1.42 | 0.084 |
| PE 18:0p_20:4;2O | 0.0079 | 0.0096 | 1.22 | 0.29 |
| PE 18:0a_20:5;O | 0.018 | 0.02 | 1.11 | 0.73 |

## References

1. Clark, S. R. et al. Characterization of platelet aminophospholipid externalization reveals fatty acids as molecular determinants that regulate coagulation. Proc Natl Acad Sci U S A 110, 5875–5880, doi:10.1073/pnas.1222419110 (2013).

2. Sivagnanam, U., Palanirajan, S. K. & Gummadi, S. N. The role of human phospholipid scramblases in apoptosis: An overview. Biochim Biophys Acta Mol Cell Res 1864, 2261–2271, doi:10.1016/j.bbamcr.2017.08.008 (2017).

3. Heemskerk, J. W., Bevers, E. M. & Lindhout, T. Platelet activation and blood coagulation. Thromb Haemost 88, 186–193 (2002).

4. Lhermusier, T., Chap, H. & Payrastre, B. Platelet membrane phospholipid asymmetry: from the characterization of a scramblase activity to the identification of an essential protein mutated in Scott syndrome. J Thromb Haemost 9, 1883–1891, doi:10.1111/j.1538-7836.2011.04478.x (2011).

5. O’Donnell, V. B., Rossjohn, J. & Wakelam, M. J. Phospholipid signaling in innate immune cells. J Clin Invest 128, 2670–2679, doi:10.1172/JCI97944 (2018).

6. van Geffen, J. P., Swieringa, F. & Heemskerk, J. W. Platelets and coagulation in thrombus formation: aberrations in the Scott syndrome. Thromb Res 141 Suppl 2, S12–16, doi:10.1016/S0049-3848(16)30355-3 (2016).

7. O’Donnell, V. B., Aldrovandi, M., Murphy, R. C. & Kronke, G. Enzymatically oxidized phospholipids assume center stage as essential regulators of innate immunity and cell death. Sci Signal 12, doi:10.1126/scisignal.aau2293 (2019).

8. Lauder, S. N. et al. Networks of enzymatically oxidized membrane lipids support calcium-dependent coagulation factor binding to maintain hemostasis. Sci Signal 10, doi:10.1126/scisignal.aan2787 (2017).

9. O’Donnell, V. B., Murphy, R. C. & Watson, S. P. Platelet lipidomics: modern day perspective on lipid discovery and characterization in platelets. Circ Res 114, 1185–1203, doi:10.1161/CIRCRESAHA.114.301597 (2014).

10. Thomas, C. P. et al. Phospholipid-esterified eicosanoids are generated in agonist-activated human platelets and enhance tissue factor-dependent thrombin generation. J Biol Chem 285, 6891–6903, doi:10.1074/jbc.M109.078428 (2010).

11. Maskrey, B. H. et al. Activated platelets and monocytes generate four hydroxyphosphatidylethanolamines via lipoxygenase. J Biol Chem 282, 20151–20163, doi:10.1074/jbc.M611776200 (2007).

12. Clark, S. R. et al. Esterified eicosanoids are acutely generated by 5-lipoxygenase in primary human neutrophils and in human and murine infection. Blood 117, 2033–2043, doi:10.1182/blood-2010-04-278887 (2011).

13. Slatter, D. A. et al. Enzymatically oxidized phospholipids restore thrombin generation in coagulation factor deficiencies. JCI Insight 3, doi:10.1172/jci.insight.98459 (2018).

14. Uderhardt, S. et al. Enzymatic lipid oxidation by eosinophils propagates coagulation, hemostasis, and thrombotic disease. J Exp Med 214, 2121–2138, doi:10.1084/jem.20161070 (2017).

15. Allen-Redpath, K. et al. Phospholipid membranes drive abdominal aortic aneurysm development through stimulating coagulation factor activity. Proc Natl Acad Sci U S A 116, 8038–8047, doi:10.1073/pnas.1814409116 (2019).

16. Anderson, J. L. et al. 2011 ACCF/AHA Focused Update Incorporated Into the ACC/AHA 2007 Guidelines for the Management of Patients With Unstable Angina/Non-ST-Elevation Myocardial Infarction: a report of the American College of Cardiology Foundation/American Heart Association Task Force on Practice Guidelines. Circulation 123, e426–579, doi:10.1161/CIR.0b013e318212bb8b (2011).

17. Armstrong, P. W. et al. Acute coronary syndromes in the GUSTO-IIb trial: prognostic insights and impact of recurrent ischemia. The GUSTO-IIb Investigators. Circulation 98, 1860–1868 (1998).

18. Hamm, C. W. et al. ESC Guidelines for the management of acute coronary syndromes in patients presenting without persistent ST-segment elevation: The Task Force for the management of acute coronary syndromes (ACS) in patients presenting without persistent ST-segment elevation of the European Society of Cardiology (ESC). Eur Heart J 32, 2999–3054, doi:10.1093/eurheartj/ehr236 (2011).

19. Liebson, P. R. & Klein, L. W. The non-Q wave myocardial infarction revisited: 10 years later. Prog Cardiovasc Dis 39, 399–444 (1997).

20. UK;, H. Key facts & figures, <https://heartuk.org.uk/press/press-kit/key-facts-figures> (2017).

21. Wilson, S. J., Newby, D. E., Dawson, D., Irving, J. & Berry, C. Duration of dual antiplatelet therapy in acute coronary syndrome. Heart 103, 573–580, doi:10.1136/heartjnl-2016-309871 (2017).

22. Mulvihill, N. T. & Foley, J. B. Inflammation in acute coronary syndromes. Heart 87, 201–204 (2002).

23. Sager, H. B. & Nahrendorf, M. Inflammation: a trigger for acute coronary syndrome. Q J Nucl Med Mol Imaging 60, 185–193 (2016).

24. Rickles, F. R., Levin, J., Hardin, J. A., Barr, C. F. & Conrad, M. E., Jr. Tissue factor generation by human mononuclear cells: effects of endotoxin and dissociation of tissue factor generation from mitogenic response. J Lab Clin Med 89, 792–803 (1977).

25. Serneri, G. G. et al. Transient intermittent lymphocyte activation is responsible for the instability of angina. Circulation 86, 790–797 (1992).

26. Mazzone, A. et al. Increased expression of neutrophil and monocyte adhesion molecules in unstable coronary artery disease. Circulation 88, 358–363 (1993).

27. Biasucci, L. M. et al. Intracellular neutrophil myeloperoxidase is reduced in unstable angina and acute myocardial infarction, but its reduction is not related to ischemia. J Am Coll Cardiol 27, 611–616 (1996).

28. Hansen, P. R. Myocardial reperfusion injury: experimental evidence and clinical relevance. Eur Heart J 16, 734–740 (1995).

29. Hedayati, T., Yadav, N. & Khanagavi, J. Non-ST-Segment Acute Coronary Syndromes. Cardiol Clin 36, 37–52, doi:10.1016/j.ccl.2017.08.003 (2018).

30. Komocsi, A., Vorobcsuk, A., Kehl, D. & Aradi, D. Use of new-generation oral anticoagulant agents in patients receiving antiplatelet therapy after an acute coronary syndrome: systematic review and meta-analysis of randomized controlled trials. Arch Intern Med 172, 1537–1545, doi:10.1001/archinternmed.2012.4026 (2012).

31. Giri, S. & Jennings, L. K. The spectrum of thrombin in acute coronary syndromes. Thromb Res 135, 782–787, doi:10.1016/j.thromres.2015.02.013 (2015).

32. Jennings, L. K. Mechanisms of platelet activation: need for new strategies to protect against platelet-mediated atherothrombosis. Thromb Haemost 102, 248–257, doi:10.1160/TH09-03-0192 (2009).

33. Moon, J. Y., Nagaraju, D., Franchi, F., Rollini, F. & Angiolillo, D. J. The role of oral anticoagulant therapy in patients with acute coronary syndrome. Ther Adv Hematol 8, 353–366, doi:10.1177/2040620717733691 (2017).

34. Neumann, F. J. et al. 2018 ESC/EACTS Guidelines on myocardial revascularization. Eur Heart J 40, 87–165, doi:10.1093/eurheartj/ehy394 (2019).

35. Hurt, L. et al. Cohort profile: HealthWise Wales. A research register and population health data platform with linkage to National Health Service data sets in Wales. BMJ Open 9, e031705, doi:10.1136/bmjopen-2019-031705 (2019).

36. Berckmans, R. J., Lacroix, R., Hau, C. M., Sturk, A. & Nieuwland, R. Extracellular vesicles and coagulation in blood from healthy humans revisited. J Extracell Vesicles 8, 1688936, doi:10.1080/20013078.2019.1688936 (2019).

37. Coumans, F. A. W. et al. Methodological Guidelines to Study Extracellular Vesicles. Circ Res 120, 1632–1648, doi:10.1161/CIRCRESAHA.117.309417 (2017).

38. Williams, C. M., Li, Y., Brown, E. & Poole, A. W. Platelet-specific deletion of SNAP23 ablates granule secretion, substantially inhibiting arterial and venous thrombosis in mice. Blood Adv 2, 3627–3636, doi:10.1182/bloodadvances.2018023291 (2018).

39. Savage, J. S. et al. Munc13-4 is critical for thrombosis through regulating release of ADP from platelets. Journal of thrombosis and haemostasis : JTH 11, 771–775, doi:10.1111/jth.12138 (2013).

40. Morgan, A. H. et al. Quantitative assays for esterified oxylipins generated by immune cells. Nat Protoc 5, 1919–1931, doi:10.1038/nprot.2010.162 (2010).

41. Slatter, D. A. et al. Mapping the Human Platelet Lipidome Reveals Cytosolic Phospholipase A2 as a Regulator of Mitochondrial Bioenergetics during Activation. Cell Metab 23, 930–944, doi:10.1016/j.cmet.2016.04.001 (2016).

42. Hong, S. H. et al. Quantitative determination of 12-hydroxyeicosatetraenoic acids by chiral liquid chromatography tandem mass spectrometry in a murine atopic dermatitis model. J Vet Sci 16, 307–315, doi:10.4142/jvs.2015.16.3.307 (2015).

43. Misheva, M. et al. Oxylipin metabolism is controlled by mitochondrial beta-oxidation during bacterial inflammation. Nat Commun 13, 139, doi:10.1038/s41467-021-27766-8 (2022).

44. Michel, M. C., Murphy, T. J. & Motulsky, H. J. New Author Guidelines for Displaying Data and Reporting Data Analysis and Statistical Methods in Experimental Biology. J Pharmacol Exp Ther 372, 136–147, doi:10.1124/jpet.119.264143 (2020).

45. von Elm, E. et al. The Strengthening the Reporting of Observational Studies in Epidemiology (STROBE) statement: guidelines for reporting observational studies. J Clin Epidemiol 61, 344–349, doi:10.1016/j.jclinepi.2007.11.008 (2008).

46. Aldrovandi, M. et al. Human platelets generate phospholipid-esterified prostaglandins via cyclooxygenase-1 that are inhibited by low dose aspirin supplementation. J Lipid Res 54, 3085–3097, doi:10.1194/jlr.M041533 (2013).

47. O’Donnell, V. B. New appreciation for an old pathway: the Lands Cycle moves into new arenas in health and disease. Biochem Soc Trans 50, 1–11, doi:10.1042/BST20210579 (2022).

48. Thuresson, E. D., Lakkides, K. M. & Smith, W. L. Different catalytically competent arrangements of arachidonic acid within the cyclooxygenase active site of prostaglandin endoperoxide H synthase-1 lead to the formation of different oxygenated products. J Biol Chem 275, 8501–8507, doi:10.1074/jbc.275.12.8501 (2000).

49. Johnsson, A. K. et al. COX-1 dependent biosynthesis of 15-hydroxyeicosatetraenoic acid in human mast cells. Biochim Biophys Acta Mol Cell Biol Lipids 1866, 158886, doi:10.1016/j.bbalip.2021.158886 (2021).

50. Powell, W. S. & Rokach, J. Biosynthesis, biological effects, and receptors of hydroxyeicosatetraenoic acids (HETEs) and oxoeicosatetraenoic acids (oxo-ETEs) derived from arachidonic acid. Biochim Biophys Acta 1851, 340–355, doi:10.1016/j.bbalip.2014.10.008 (2015).

51. Mazaleuskaya, L. L. et al. Analysis of HETEs in human whole blood by chiral UHPLC-ECAPCI/HRMS. J Lipid Res 59, 564–575, doi:10.1194/jlr.D081414 (2018).

52. Siebert, M., Krieg, P., Lehmann, W. D., Marks, F. & Furstenberger, G. Enzymic characterization of epidermis-derived 12-lipoxygenase isoenzymes. Biochem J 355, 97–104, doi:10.1042/0264-6021:3550097 (2001).

53. Protty, M. B., Jenkins, P. V., Collins, P. W. & O’Donnell, V. B. The role of procoagulant phospholipids on the surface of circulating blood cells in thrombosis and haemostasis. Open Biol 12, 210318, doi:10.1098/rsob.210318 (2022).

54. Sun, F. F., McGuire, J. C. & Metzler, C. M. The effect of substrate availability on the metabolism of arachidonic acid in human platelets. Prog Lipid Res 20, 275–278, doi:10.1016/0163-7827(81)90054-0 (1981).

55. Hua, V. M. et al. Necrotic platelets provide a procoagulant surface during thrombosis. Blood 126, 2852–2862, doi:10.1182/blood-2015-08-663005 (2015).

56. Agbani, E. O. & Poole, A. W. Procoagulant platelets: generation, function, and therapeutic targeting in thrombosis. Blood 130, 2171–2179, doi:10.1182/blood-2017-05-787259 (2017).

57. Prodan, C. I., Joseph, P. M., Vincent, A. S. & Dale, G. L. Coated-platelet levels are influenced by smoking, aspirin, and selective serotonin reuptake inhibitors. J Thromb Haemost 5, 2149–2151, doi:10.1111/j.1538-7836.2007.02691.x (2007).

58. Braun, O. O. et al. Greater reduction of platelet activation markers and platelet-monocyte aggregates by prasugrel compared to clopidogrel in stable coronary artery disease. Thromb Haemost 100, 626–633 (2008).

59. Klinkhardt, U. et al. Clopidogrel but not aspirin reduces P-selectin expression and formation of platelet-leukocyte aggregates in patients with atherosclerotic vascular disease. Clin Pharmacol Ther 73, 232–241, doi:10.1067/mcp.2003.13 (2003).

60. Blankenberg, S. & Yusuf, S. The inflammatory hypothesis: any progress in risk stratification and therapeutic targets? Circulation 114, 1557–1560, doi:10.1161/CIRCULATIONAHA.106.652081 (2006).

61. Libby, P. et al. Inflammation, Immunity, and Infection in Atherothrombosis: JACC Review Topic of the Week. J Am Coll Cardiol 72, 2071–2081, doi:10.1016/j.jacc.2018.08.1043 (2018).

62. Libby, P., Nahrendorf, M. & Swirski, F. K. Leukocytes Link Local and Systemic Inflammation in Ischemic Cardiovascular Disease: An Expanded “Cardiovascular Continuum”. J Am Coll Cardiol 67, 1091–1103, doi:10.1016/j.jacc.2015.12.048 (2016).

63. Otahbachi, M. et al. Gender differences in platelet aggregation in healthy individuals. J Thromb Thrombolysis 30, 184–191, doi:10.1007/s11239-009-0436-x (2010).

64. Ranucci, M. et al. Gender-based differences in platelet function and platelet reactivity to P2Y12 inhibitors. PLoS One 14, e0225771, doi:10.1371/journal.pone.0225771 (2019).

65. Haque, S. F. et al. Sex difference in platelet aggregation detected by new aggregometry using light scattering. Endocr J 48, 33–41, doi:10.1507/endocrj.48.33 (2001).

66. Naruko, T. et al. Neutrophil infiltration of culprit lesions in acute coronary syndromes. Circulation 106, 2894–2900, doi:10.1161/01.cir.0000042674.89762.20 (2002).

67. Waddington, E., Sienuarine, K., Puddey, I. & Croft, K. Identification and quantitation of unique fatty acid oxidation products in human atherosclerotic plaque using high-performance liquid chromatography. Anal Biochem 292, 234–244, doi:10.1006/abio.2001.5075 (2001).

68. Kuhn, H., Heydeck, D., Hugou, I. & Gniwotta, C. In vivo action of 15-lipoxygenase in early stages of human atherogenesis. J Clin Invest 99, 888–893, doi:10.1172/JCI119253 (1997).

69. Asada, Y., Yamashita, A., Sato, Y. & Hatakeyama, K. Pathophysiology of atherothrombosis: Mechanisms of thrombus formation on disrupted atherosclerotic plaques. Pathol Int, doi:10.1111/pin.12921 (2020).

70. Mukhopadhyay, S. et al. Fibrinolysis and Inflammation in Venous Thrombus Resolution. Front Immunol 10, 1348, doi:10.3389/fimmu.2019.01348 (2019).

71. Goto, Y. et al. Degradation of phospholipid molecular species during experimental cerebral ischemia in rats. Stroke 19, 728–735, doi:10.1161/01.str.19.6.728 (1988).

72. Ong, S. B. et al. Inflammation following acute myocardial infarction: Multiple players, dynamic roles, and novel therapeutic opportunities. Pharmacol Ther 186, 73–87, doi:10.1016/j.pharmthera.2018.01.001 (2018).

73. Ittaman, S. V., VanWormer, J. J. & Rezkalla, S. H. The role of aspirin in the prevention of cardiovascular disease. Clin Med Res 12, 147–154, doi:10.3121/cmr.2013.1197 (2014).

74. Protty, M. B., Wilkins, S. J., Hoskins, H. C., Dawood, B. B. & Hayes, J. Prescribing patterns of oral antiplatelets in Wales: evolving trends from 2005 to 2016. Future Cardiol 14, 277–282, doi:10.2217/fca-2018-0003 (2018).

75. Vane, J. R. & Botting, R. M. The mechanism of action of aspirin. Thromb Res 110, 255–258, doi:10.1016/s0049-3848(03)00379-7 (2003).

76. Koupenova, M., Kehrel, B. E., Corkrey, H. A. & Freedman, J. E. Thrombosis and platelets: an update. Eur Heart J 38, 785–791, doi:10.1093/eurheartj/ehw550 (2017).

